# Characterization of sleep apnea among a sample of adults from Samoa

**DOI:** 10.1101/2023.11.16.23298644

**Authors:** Lacey W. Heinsberg, Alysa Pomer, Brian E. Cade, Jenna C. Carlson, Take Naseri, Muagututia Sefuiva Reupena, Satupa’itea Viali, Daniel E. Weeks, Stephen T. McGarvey, Susan Redline, Nicola L. Hawley

**Author notes:** Corresponding author: Lacey W. Heinsberg, PhD, RN, Department of Human Genetics School of Public Health University of Pittsburgh, Public Health 3121G 130 De Soto Street Pittsburgh, PA 15261, |. Authors Email addresses: Lacey W. Heinsberg, Alysa Pomer, Brian E. Cade, Jenna C. Carlson, Take Naseri, Muagututia Sefuiva Reupena, Satupa’itea Viali, Daniel E. Weeks, Stephen McGarvey, Susan Redline, Nicola L. Hawley.

## Abstract

Sleep apnea is a public health concern around the world, but little research has been dedicated to examining this issue in low- and middle-income countries, including Samoa. Using data collected through the *Soifua Manuia* (“Good Health”) study, which aimed to investigate the impact of the body mass index (BMI)-associated genetic variant rs373863828 in CREB3 Regulatory Factor (*CREBRF*) on metabolic traits in Samoan adults, we examined the sample prevalence and characteristics of sleep apnea using data collected with a validated home sleep apnea device (WatchPAT, Itamar). A total of 330 participants (sampled to overrepresent the obesity-risk allele of interest) had sleep data available. Participants (53.3% female) had a mean (SD) age of 52.0 (9.9) years and BMI of 35.5 (7.5) kg/m^2^ and 36.3% of the sample had type 2 diabetes. Based on the 3% and 4% apnea hypopnea indices (AHI) and the 4% oxygen desaturation index (ODI), descriptive analyses revealed that many participants had potentially actionable sleep apnea defined as >5 events/hr (87.9%, 68.5%, and 71.2%, respectively) or clinically actionable sleep apnea defined as ≥15 events/hr (54.9%, 31.5%, and 34.5%, respectively). Sleep apnea was more severe in men; for example, clinically actionable sleep apnea (≥15) based on the AHI 3% definition was observed in 61.7% of men and 48.9% of women. Correction for non-representational sampling related to the *CREBRF* obesity-risk allele resulted in only slightly lower estimates. Across the AHI 3%, AHI 4%, and ODI 4%, multiple linear regression revealed associations between a greater number of events/hr and higher age, male sex, higher body mass index, higher abdominal-hip circumference ratio, and geographic region of residence. Our study identified a much higher frequency of sleep apnea in Samoa compared with published data from other studies, but similar predictors. Continued research addressing generalizability of these findings, as well as a specific focus on diagnosis and affordable and equitable access to treatment, is needed to alleviate the burden of sleep apnea in Samoa and around the world.

## 1. INTRODUCTION

Sleep apnea is a prevalent disorder characterized by recurring upper airway obstruction, resulting in reduced oxygen levels and fragmented sleep patterns. It has far-reaching negative consequences, encompassing compromised physical health, impaired cognitive function, and diminished overall quality of life.^1^ Globally, sleep apnea is estimated to affect as many as 1 billion individuals, making it a significant public health concern.^2^ While sleep apnea has been extensively studied in high-income countries, there remains a dearth of high-quality sleep data from low- and middle-income countries^3^, including Samoa. This research gap is particularly concerning, given that Samoa has among the highest global prevalence of obesity^4,5^, one of the most influential known risk factors for sleep apnea.

Beyond Samoa specifically, there is a dearth of research examining sleep health in the Pacific Islander population group broadly. Among the existing studies, it is consistently evident that sleep health is a significant concern among this population. For example, Native Hawaiians and Pacific Islanders in the United States have been reported to experience insufficient sleep duration and elevated daytime sleepiness.^6,7^ Moreover, a retrospective chart review of Native Hawaiians and Pacific Islander patients who visited a sleep center in Utah found that the participants not only exhibited high rates of sleep apnea (which was expected given the sleep center setting), but that sleep apnea data were skewed toward greater disorder severity and that there was generally low uptake/adherence to prescribed sleep apnea treatment.^8^ Further, a sleep apnea study in New Zealand found that Māori individuals had a higher prevalence of sleep apnea compared to non-Māori individuals.^9^ It’s worth noting that sleep apnea is an independent risk factor for all-cause mortality^10^ but, fortunately, there are treatment interventions available that can reduce this risk. For example, Continuous Positive Airway Pressure (CPAP) can be effective in not only improving sleep quality,^11^ but also enhancing glycemic control, reducing insulin resistance, improving fasting glucose, lowering hemoglobin A1c levels, and decreasing blood pressure.^12,13^ Given the existence of effective interventions for sleep apnea,^14^ prioritizing sleep as an integral component of holistic health should be a paramount concern to address inequalities in health seen in Pacific Islander populations.

These findings, paired with our prior work which highlights a high sample prevalence of excessive daytime sleepiness in Samoa,^15^ support the need to characterize the underlying sleep pathology – specifically the burden of sleep apnea – as it could shed light on the unique challenges and potential preventive measures that could improve the well-being of this population. Therefore, the purpose of this study was to examine the prevalence and characteristics of sleep apnea in a sample of adults from Samoa.

## 2. MATERIALS AND METHODS

### 2.1 STUDY OVERVIEW

This was a secondary analysis of data collected via the 2017-2019 *Soifua Manuia* (“Good Health”) study which aimed to investigate the impact of the CREB3 Regulatory Factor (*CREBRF*) genetic variant rs373863828 on metabolic traits in Samoan adults.^16^ The variant’s minor allele (A) is paradoxically associated with increased body mass index (BMI), but lower odds of type 2 diabetes mellitus (T2DM) in Pacific Islander individuals.^17^ The overall sample included 519 participants from ‘Upolu, Samoa, with intentional oversampling of the *CREBRF* obesity-risk A allele at an approximate GG:AG:AA genotype ratio of 2:2:1 (compared with the expected population ratio of 8.2:5.7:1). All participants completed extensive physical assessments and comprehensive behavioral questionnaires. Objective sleep data were collected in a subset of participants as the assessment was added several months into the recruitment period. The study protocol and recruitment procedures are detailed elsewhere.^16^ The study received approval from ethical boards at Yale University, Brown University, the University of Pittsburgh, Mass General Brigham, and the Samoa Ministry of Health. All participants gave written informed consent.

### 2.2 PHENOTYPE DATA

Objective sleep data were collected using WatchPAT TM 200 Unified (Itamar Medical Ltd.) devices worn on the nondominant wrist. These highly accurate and clinically reliable devices measure peripheral arterial tone, heart rate, oxygen saturation, actigraphy, snoring levels, and body position via three points of contact (wrist, finger, and chest). Of note, the WatchPAT apnea-hypopnea and oxygen desaturation indices demonstrate an impressive 93% to 99% correlation when compared to the same metrics collected using the gold standard polysomnography.^18^ Data were reviewed and cleaned by trained sleep research polysomnologists (Methods S1). This report primarily focuses on three clinical indicators of sleep apnea: the apnea-hypopnea index (AHI) using both 3% and 4% oxygen desaturation cutoffs, and the oxygen desaturation index (ODI) using a 4% oxygen desaturation cutoff. The AHI quantifies the number of apneas (pauses in breathing) and hypopneas (partial reductions in breathing) associated with a ≥3% (AHI 3%) or ≥4% (AHI 4%) reduction in blood oxygen levels per hour of sleep which is estimated by the WatchPAT device using changes in peripheral arterial tonometry and changes in heart rate and oxygen saturation. The ODI measures the frequency of ≥4% oxygen desaturation events per hour (events/hr) of sleep irrespective of apneas/hypopneas. Secondary measures included the AHI 3%, AHI 4%, and ODI 4% across supine and non-supine sleep positions and rapid eye movement (REM) and non-REM estimates of sleep states using the devices’ proprietary algorithm^19^ as well as percent time spent below 90% oxygen saturation (T90). Sleep apnea measures were examined as continuous events/hr and collapsed based on clinically accepted sleep apnea categories by number of events/hr (none/minimal: <5; mild: 5 to <15; moderate: 15 to < 30; and severe: ≥30).^20^ To further support the descriptive nature of this paper, sleep apnea measures were further collapsed into categories of “potentially actionable” (>5 events/hr) and “clinically actionable” (≥15 events/hr). Of note, the inclusion of the >5 events/hr category, although not typically considered of major concern in the absence of symptoms, offers important insight into the characterization of potential sleep apnea in this population. Participants provided data about the night of the WatchPAT assessment related to sleep surface (e.g., mattress on a raised bed, mat on the floor, etc.), the number of people sharing the sleep surface, and the number of people sharing the room. The assessment season was inferred based on assessment date.

AHI and ODI distributions were examined by participant characteristics that have been associated with sleep apnea in other populations^21^ and were available in the existing data. Age and sex were self-reported by participants. Weight and height were measured in duplicate using a digital scale (Tanita HD 351; Tanita Corporation of America) and portable stadiometer (SECA 213, Seca GmbH & Co), and BMI was computed from the averaged values. BMI was treated as continuous and categorical variables using Polynesian cutoffs for underweight (<18kg/m^2^), ‘normal’ (18 to <26 kg/m^2^), overweight (26 to 32 kg/m^2^), and obesity (>32 kg/m^2^). Abdominal and hip circumference were measured using a standard tape measure (SECA 201, SECA, Hamburg, Germany) and abdominal-hip ratio was computed. *CREBRF* rs373863828 genotype data were generated via TaqMan real-time PCR (Applied Biosystems).^17^ T2DM status was determined based on any of the following: current use of diabetes medication; Hemoglobin A1c (HbA1c) (5.7-6.4%, pre-diabetes; >6.4%, diabetes); fasting blood glucose (FBG; 100-125, pre-diabetes; ≥126, diabetes); and/or oral glucose tolerance testing based on the American Diabetes Association criteria.^22^ Blood pressure (BP) was measured in triplicate and the second and third values were averaged; hypertension was defined as hypertension medication use or an average systolic BP ≥140 or average diastolic BP ≥90 mmHg. Participants self-reported asthma diagnoses. Behavioral and social data encompassed smoking habits (current use of cigarettes, tobacco, or pipe; yes/no), alcohol consumption in the past 12 months (yes/no), and socioeconomic resources quantified using a material lifestyle score^23^ which sums the number of items owned from an 18-item household inventory (refrigerator, freezer, portable stereo/MP3 player, etc.); for participants with a low level (i.e., <35%) of missing information across the questionnaire, imputation was used to fill in missing data.^15^ Moderate-vigorous physical activity data (average minutes per day) were collected using Actigraph GT3X+ accelerometer-based devices (ActiGraph Corporation) as detailed elsewhere.^24^

### 2.3 STATISTICAL ANALYSES

Statistical and descriptive analyses were conducted using R version 4.1.2.^25^ Data were described using means, standard deviations (SD), medians, and interquartile ranges (IQR) for continuous variables and frequency counts and percentages for categorical variables. Categorical sleep apnea sample prevalence was calculated across the three primary sleep apnea measures (AHI 3%, AHI 4%, and ODI 4%). To evaluate the influence of non-representational sampling and to obtain more accurate estimates of sleep apnea prevalence that might be seen in a random sample of the population, we performed a correction for *CREBRF* obesity-risk allele enrichment (Code S1). Specifically, we calculated genotype-specific weights by comparing the expected genotype frequencies in the general population with the observed genotype frequencies in our sample. Similar to inverse probability weighting, we then used the genotype-specific weights to adjust the counts of categorical sleep apnea severity (none, mild, moderate, severe).

To better understand AHI and ODI data distributions across the participant factors described above, various visualizations were employed including stacked bar, sina with violin, rain, scatter, and mosaic plots. Associations between sleep measures and participant factors were formally tested using Spearman correlation coefficients and Wilcoxon or Kruskal-Wallis rank sum tests depending on the predictor variable type. Finally, multiple linear regression was used to examine associations of square root transformed (given data skewness) sleep apnea measures and *a priori* selected participant characteristics of age, sex, *CREBRF* genotype, BMI, abdominal-hip ratio, T2DM, hypertension, asthma, census region, relationship status, and cigarette and alcohol use. Unstandardized regression estimates, 95% confidence intervals (CI), and p-values were reported, and model assessment was performed using residual analysis and influence diagnostics. Across all analyses, p-values < 0.05 were considered statistically significant.

## 3. RESULTS

The overall sample with acceptable WatchPAT data available consisted of 330 participants (53.3% female) with a mean (±SD) age of 52.0 (±9.9) years and BMI of 35.5 (±7.5) kg/m^2^ (Table 1). No statistically significant differences were observed between the WatchPAT sample and the larger parent sample (Table S1).

**Table 1.**
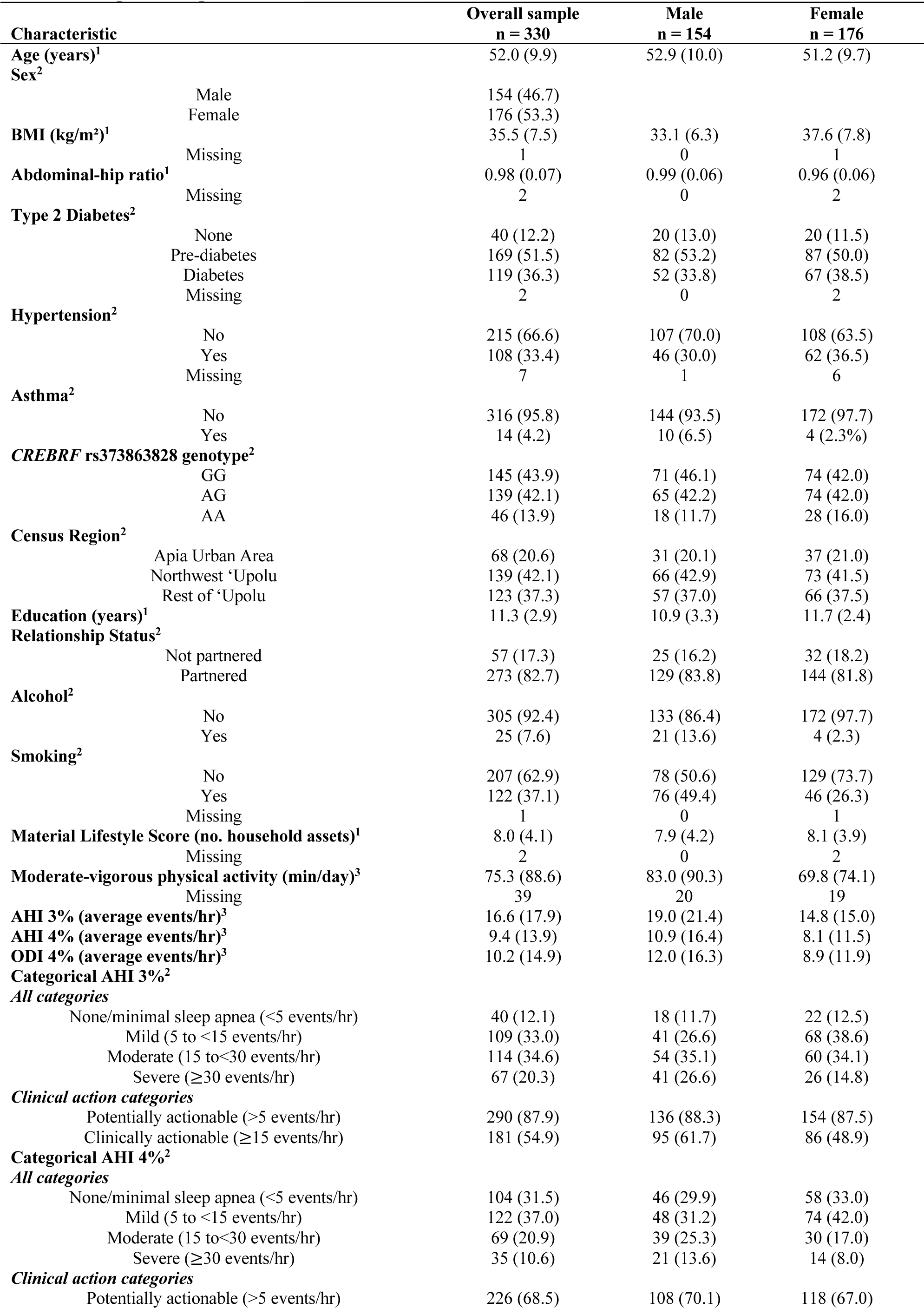
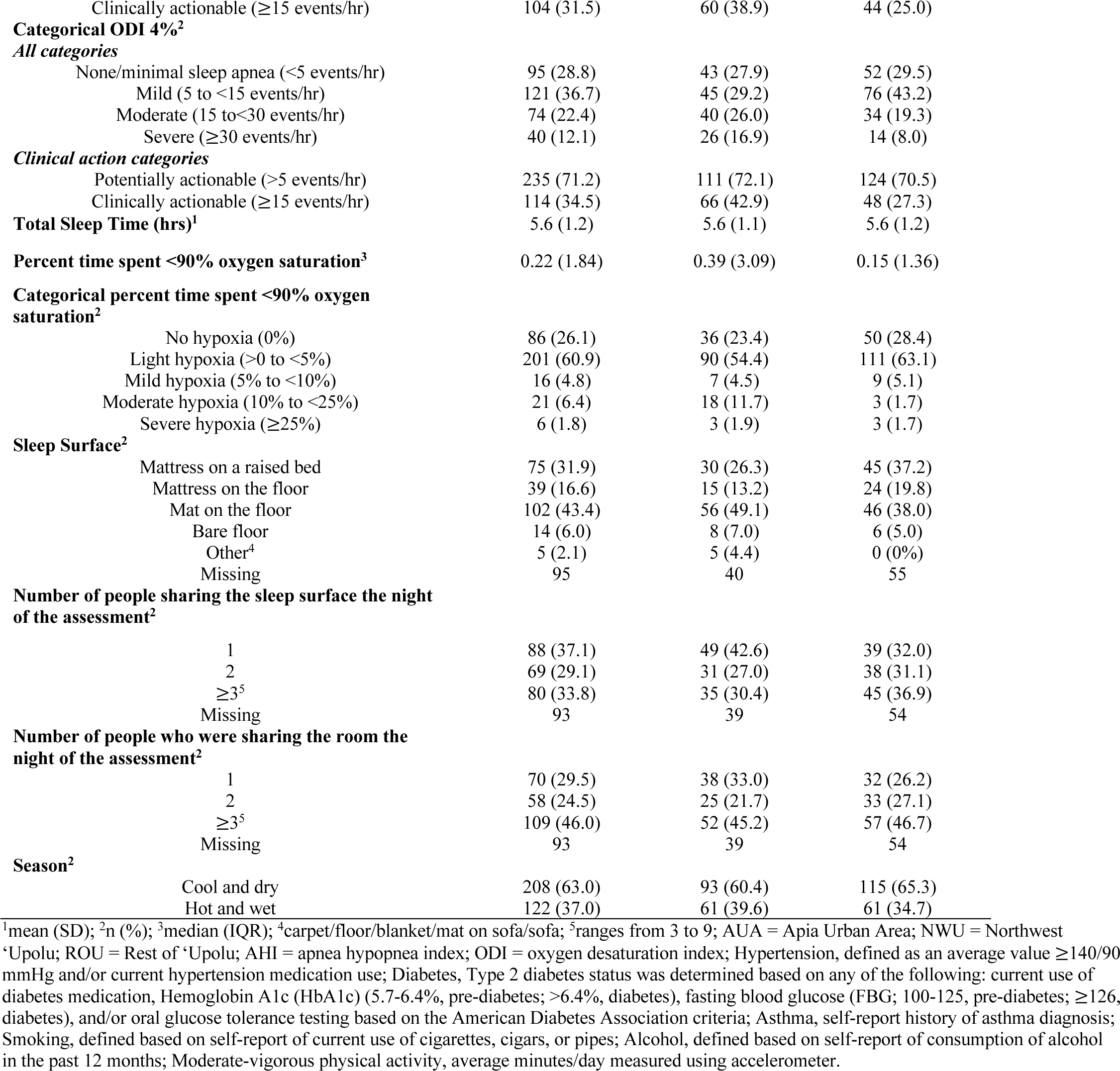
Sample description and characteristics.

| Characteristic |  | Overall sample<br>n = 330 | Male<br>n = 154 | Female<br>n = 176 |
| --- | --- | --- | --- | --- |
| Age (years)<sup>1</sup> |  | 52.0 (9.9) | 52.9 (10.0) | 51.2 (9.7) |
| Sex<sup>2</sup> |  |  |  |  |
|  | Male | 154 (46.7) |  |  |
|  | Female | 176 (53.3) |  |  |
| BMI (kg/m<sup>2</sup>)<sup>1</sup> |  | 35.5 (7.5) | 33.1 (6.3) | 37.6 (7.8) |
|  | Missing | 1 | 0 | 1 |
| Abdominal-hip ratio<sup>1</sup> |  | 0.98 (0.07) | 0.99 (0.06) | 0.96 (0.06) |
|  | Missing | 2 | 0 | 2 |
| Type 2 Diabetes<sup>2</sup> |  |  |  |  |
|  | None | 40 (12.2) | 20 (13.0) | 20 (11.5) |
|  | Pre-diabetes | 169 (51.5) | 82 (53.2) | 87 (50.0) |
|  | Diabetes | 119 (36.3) | 52 (33.8) | 67 (38.5) |
|  | Missing | 2 | 0 | 2 |
| Hypertension<sup>2</sup> |  |  |  |  |
|  | No | 215 (66.6) | 107 (70.0) | 108 (63.5) |
|  | Yes | 108 (33.4) | 46 (30.0) | 62 (36.5) |
|  | Missing | 7 | 1 | 6 |
| Asthma<sup>2</sup> |  |  |  |  |
|  | No | 316 (95.8) | 144 (93.5) | 172 (97.7) |
|  | Yes | 14 (4.2) | 10 (6.5) | 4 (2.3%) |
| CREBRF rs373863828 genotype<sup>2</sup> |  |  |  |  |
|  | GG | 145 (43.9) | 71 (46.1) | 74 (42.0) |
|  | AG | 139 (42.1) | 65 (42.2) | 74 (42.0) |
|  | AA | 46 (13.9) | 18 (11.7) | 28 (16.0) |
| Census Region<sup>2</sup> |  |  |  |  |
|  | Apia Urban Area | 68 (20.6) | 31 (20.1) | 37 (21.0) |
|  | Northwest 'Upolu | 139 (42.1) | 66 (42.9) | 73 (41.5) |
|  | Rest of 'Upolu | 123 (37.3) | 57 (37.0) | 66 (37.5) |
| Education (years)<sup>1</sup> |  | 11.3 (2.9) | 10.9 (3.3) | 11.7 (2.4) |
| Relationship Status<sup>2</sup> |  |  |  |  |
|  | Not partnered | 57 (17.3) | 25 (16.2) | 32 (18.2) |
|  | Partnered | 273 (82.7) | 129 (83.8) | 144 (81.8) |
| Alcohol<sup>2</sup> |  |  |  |  |
|  | No | 305 (92.4) | 133 (86.4) | 172 (97.7) |
|  | Yes | 25 (7.6) | 21 (13.6) | 4 (2.3) |
| Smoking<sup>2</sup> |  |  |  |  |
|  | No | 207 (62.9) | 78 (50.6) | 129 (73.7) |
|  | Yes | 122 (37.1) | 76 (49.4) | 46 (26.3) |
|  | Missing | 1 | 0 | 1 |
| Material Lifestyle Score (no. household assets)<sup>1</sup> |  | 8.0 (4.1) | 7.9 (4.2) | 8.1 (3.9) |
|  | Missing | 2 | 0 | 2 |
| Moderate-vigorous physical activity (min/day)<sup>3</sup> |  | 75.3 (88.6) | 83.0 (90.3) | 69.8 (74.1) |
|  | Missing | 39 | 20 | 19 |
| AHI 3% (average events/hr)<sup>3</sup> |  | 16.6 (17.9) | 19.0 (21.4) | 14.8 (15.0) |
| AHI 4% (average events/hr)<sup>3</sup> |  | 9.4 (13.9) | 10.9 (16.4) | 8.1 (11.5) |
| ODI 4% (average events/hr)<sup>3</sup> |  | 10.2 (14.9) | 12.0 (16.3) | 8.9 (11.9) |
| Categorical AHI 3%<sup>2</sup> |  |  |  |  |
| *All categories* |  |  |  |  |
|  | None/minimal sleep apnea (<5 events/hr) | 40 (12.1) | 18 (11.7) | 22 (12.5) |
|  | Mild (5 to <15 events/hr) | 109 (33.0) | 41 (26.6) | 68 (38.6) |
|  | Moderate (15 to<30 events/hr) | 114 (34.6) | 54 (35.1) | 60 (34.1) |
|  | Severe (≥30 events/hr) | 67 (20.3) | 41 (26.6) | 26 (14.8) |
| *Clinical action categories* |  |  |  |  |
|  | Potentially actionable (>5 events/hr) | 290 (87.9) | 136 (88.3) | 154 (87.5) |
|  | Clinically actionable (≥15 events/hr) | 181 (54.9) | 95 (61.7) | 86 (48.9) |
| Categorical AHI 4%<sup>2</sup> |  |  |  |  |
| *All categories* |  |  |  |  |
|  | None/minimal sleep apnea (<5 events/hr) | 104 (31.5) | 46 (29.9) | 58 (33.0) |
|  | Mild (5 to <15 events/hr) | 122 (37.0) | 48 (31.2) | 74 (42.0) |
|  | Moderate (15 to<30 events/hr) | 69 (20.9) | 39 (25.3) | 30 (17.0) |
|  | Severe (≥30 events/hr) | 35 (10.6) | 21 (13.6) | 14 (8.0) |
| *Clinical action categories* |  |  |  |  |
|  | Potentially actionable (>5 events/hr) | 226 (68.5) | 108 (70.1) | 118 (67.0) |

|  |  |  |  |
| --- | --- | --- | --- |
| Clinically actionable ($\geq 15$ events/hr) | 104 (31.5) | 60 (38.9) | 44 (25.0) |
| **Categorical ODI 4%<sup>2</sup>** |  |  |  |
| *All categories* |  |  |  |
| None/minimal sleep apnea (<5 events/hr) | 95 (28.8) | 43 (27.9) | 52 (29.5) |
| Mild (5 to <15 events/hr) | 121 (36.7) | 45 (29.2) | 76 (43.2) |
| Moderate (15 to <30 events/hr) | 74 (22.4) | 40 (26.0) | 34 (19.3) |
| Severe ($\geq 30$ events/hr) | 40 (12.1) | 26 (16.9) | 14 (8.0) |
| *Clinical action categories* |  |  |  |
| Potentially actionable (>5 events/hr) | 235 (71.2) | 111 (72.1) | 124 (70.5) |
| Clinically actionable ($\geq 15$ events/hr) | 114 (34.5) | 66 (42.9) | 48 (27.3) |
| **Total Sleep Time (hrs)<sup>1</sup>** | 5.6 (1.2) | 5.6 (1.1) | 5.6 (1.2) |
| **Percent time spent <90% oxygen saturation<sup>3</sup>** | 0.22 (1.84) | 0.39 (3.09) | 0.15 (1.36) |
| **Categorical percent time spent <90% oxygen saturation<sup>2</sup>** |  |  |  |
| No hypoxia (0%) | 86 (26.1) | 36 (23.4) | 50 (28.4) |
| Light hypoxia (>0 to <5%) | 201 (60.9) | 90 (54.4) | 111 (63.1) |
| Mild hypoxia (5% to <10%) | 16 (4.8) | 7 (4.5) | 9 (5.1) |
| Moderate hypoxia (10% to <25%) | 21 (6.4) | 18 (11.7) | 3 (1.7) |
| Severe hypoxia ($\geq 25\%$) | 6 (1.8) | 3 (1.9) | 3 (1.7) |
| **Sleep Surface<sup>2</sup>** |  |  |  |
| Mattress on a raised bed | 75 (31.9) | 30 (26.3) | 45 (37.2) |
| Mattress on the floor | 39 (16.6) | 15 (13.2) | 24 (19.8) |
| Mat on the floor | 102 (43.4) | 56 (49.1) | 46 (38.0) |
| Bare floor | 14 (6.0) | 8 (7.0) | 6 (5.0) |
| Other<sup>4</sup> | 5 (2.1) | 5 (4.4) | 0 (0%) |
| Missing | 95 | 40 | 55 |
| **Number of people sharing the sleep surface the night of the assessment<sup>2</sup>** |  |  |  |
| 1 | 88 (37.1) | 49 (42.6) | 39 (32.0) |
| 2 | 69 (29.1) | 31 (27.0) | 38 (31.1) |
| $\geq 3^5$ | 80 (33.8) | 35 (30.4) | 45 (36.9) |
| Missing | 93 | 39 | 54 |
| **Number of people who were sharing the room the night of the assessment<sup>2</sup>** |  |  |  |
| 1 | 70 (29.5) | 38 (33.0) | 32 (26.2) |
| 2 | 58 (24.5) | 25 (21.7) | 33 (27.1) |
| $\geq 3^5$ | 109 (46.0) | 52 (45.2) | 57 (46.7) |
| Missing | 93 | 39 | 54 |
| **Season<sup>2</sup>** |  |  |  |
| Cool and dry | 208 (63.0) | 93 (60.4) | 115 (65.3) |
| Hot and wet | 122 (37.0) | 61 (39.6) | 61 (34.7) |

<sup>1</sup>mean (SD); <sup>2</sup>n (%); <sup>3</sup>median (IQR); <sup>4</sup>carpet/floor/blanket/mat on sofa/sofa; <sup>5</sup>ranges from 3 to 9; AUA = Apia Urban Area; NWU = Northwest 'Upolu; ROU = Rest of 'Upolu; AHI = apnea hypopnea index; ODI = oxygen desaturation index; Hypertension, defined as an average value $\geq 140/90$ mmHg and/or current hypertension medication use; Diabetes, Type 2 diabetes status was determined based on any of the following: current use of diabetes medication, Hemoglobin A1c (HbA1c) (5.7-6.4%, pre-diabetes; $>6.4\%$ , diabetes), fasting blood glucose (FBG; 100-125, pre-diabetes; $\geq 126$ , diabetes), and/or oral glucose tolerance testing based on the American Diabetes Association criteria; Asthma, self-report history of asthma diagnosis; Smoking, defined based on self-report of current use of cigarettes, cigars, or pipes; Alcohol, defined based on self-report of consumption of alcohol in the past 12 months; Moderate-vigorous physical activity, average minutes/day measured using accelerometer.

The majority of the sample, 87.9%, 68.5%, and 71.2%, had potentially actionable sleep apnea (>5 events/hr) while 54.9%, 31.5%, and 34.5% had clinically actionable sleep apnea (≥15 events/hr) based on the AHI 3%, AHI 4%, and ODI 4%, respectively (Table 1). Focusing on clinically actionable sleep apnea (≥15 events/hr), rates were higher among men (61.7%, 38.9%, and 42.9%) vs. women (48.9%, 25.0%, and 27.3%) across AHI 3%, AHI 4%, and ODI 4%, respectively. Higher AHI and ODI scores were observed in supine (vs. non-supine) sleep positions and REM (vs. non-REM) sleep cycles (Table S2). Empirical correction to obtain prevalence estimates that better reflect the sleep apnea severity distribution that might be seen in the broader population, accounting for the overrepresentation of the *CREBRF* obesity-risk allele associated BMI inflation in the sample, revealed only small shifts in sleep apnea categories across the three measures (Table S3). Specifically, percentages in each sleep apnea category had a maximum shift of 1.6%, and the estimated percentage of individuals with clinically actionable sleep apnea (>5 events/hr) dropped by only 0.1% to 1.2% across the three measures, so *CREBRF*-adjusted prevalences were very similar to the raw frequencies shown in Table 1. Despite the high AHI/ODI levels, 87% of the sample had no hypoxia (26.1% had a T90 of 0%) or light hypoxia (60.9% of the sample had a T90 of >0% to <5%) (Table 1). Greater hypoxia was observed in individuals with more severe sleep apnea (Table S4).

Stacked bar plots depict the distribution of categorical sleep apnea categories across a variety of different factors (Figure 1). Consistent with the observed distributions seen in Figure 1, more detailed examination of the relationships between continuous (i.e., without collapsing) AHI/ODI data and categorical (Figures S1-S3) and continuous (Figure S4) participant factors showed that a greater number of events/hr was associated with increased age, BMI, abdominal-hip ratio, and number of household assets; lower physical activity levels; the presence of hypertension; and *CREBRF* genotype. Exploratory analyses revealed that the association between sleep apnea measures and *CREBRF* genotype persisted when controlling for age, sex, and T2DM status but did not persist when controlling for age, sex, and BMI (Table S5). Exploratory mosaic plots (Figures S5-S8) depicting the proportional distribution of sleep apnea categories across a variety of factors support joint effects of sex, age, and BMI on sleep apnea severity.

**Figure 1.**
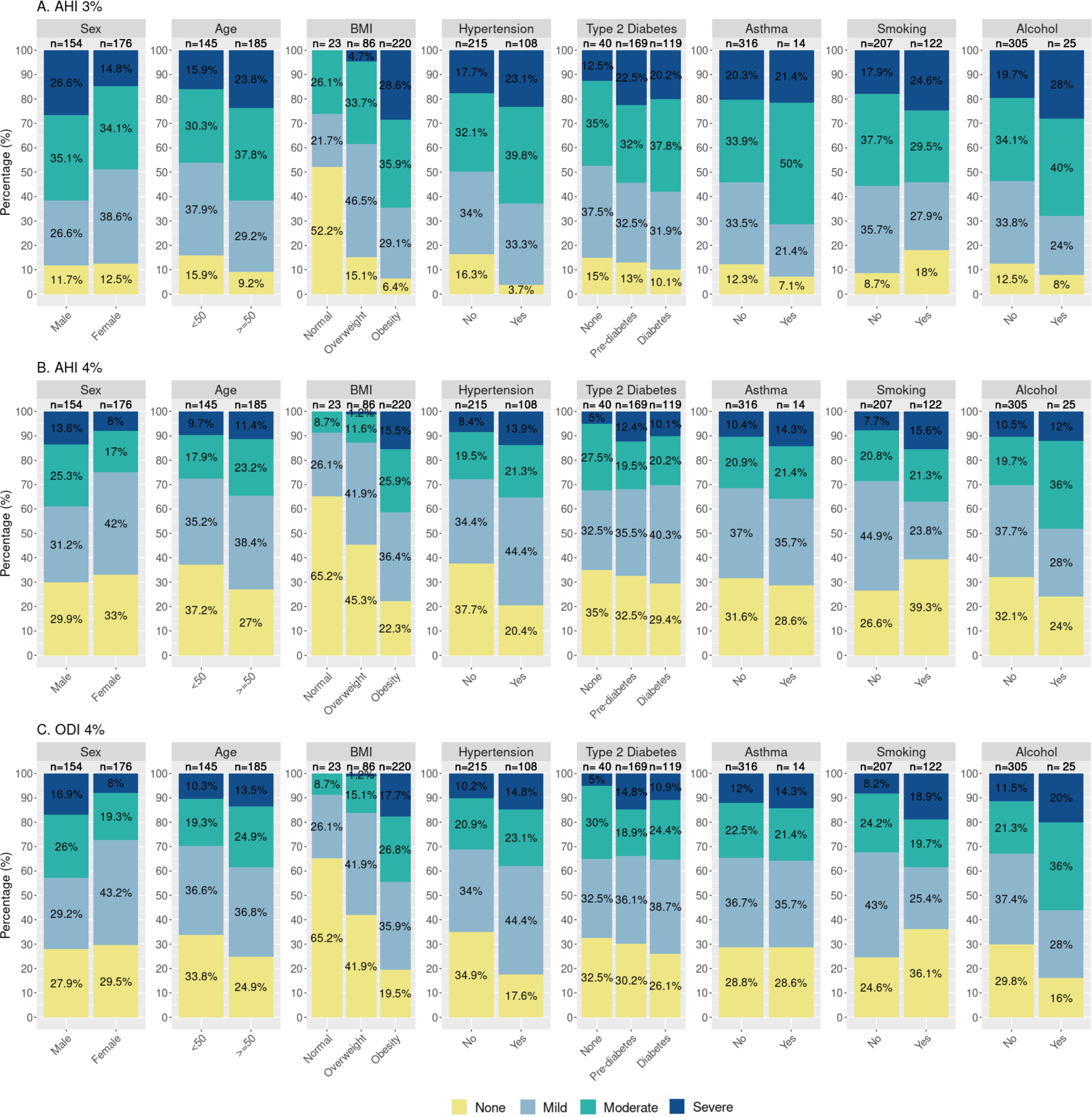
Stacked bar plots of sleep apnea measures by participant characteristics. AHI = apnea hypopnea index; ODI = oxygen desaturation index; None = <5 events/hr; Mild = 5-14.9 events/hr; Moderate = 15-29.9 events/hr; Severe = ≥30 events/hr. Age (years); BMI (kg/m^2^), ‘Normal’ = 18-25.99 kg/m^2^, Overweight = 26-32 kg/m^2^, Obesity = >32 kg/m^2^; Hypertension, defined as an average value ≥140/90 mmHg and/or current hypertension medication use; Diabetes, Type 2 diabetes status was determined based on any of the following: current use of diabetes medication; Hemoglobin A1c (HbA1c) (5.7-6.4%, pre-diabetes; >6.4%, diabetes); fasting blood glucose (FBG; 100-125, pre-diabetes; ≥126, diabetes); and/or oral glucose tolerance testing based on the American Diabetes Association criteria; Asthma, self-report history of asthma diagnosis; Smoking, defined based on self-report of current use of cigarettes, cigars, or pipes; Alcohol, defined based on self-report of consumption of alcohol in the past 12 months; see Figures S1 and S2 for additional descriptive plots.

Finally, the results of linear regression examining associations of square root-transformed sleep apnea with *a priori* selected participant characteristics (age, sex, *CREBRF* genotype, BMI, abdominal-hip circumference ratio, T2DM status, hypertension status, asthma status, census region of residence, relationship status, socioeconomic resources, education, cigarette use, and alcohol use) are presented (Table 2). We observed associations between higher AHI/ODI levels and higher age (*β*=0.03, p=0.001 to 0.002); male sex (*β*=0.884 to 0.979, p=1.52E-07 to 7.99E-08); higher BMI (*β*=0.124 to 0.136, p=6.16E-21 to 9.35E-19); higher abdominal-hip circumference ratio (*β*=2.875 to 2.923, p=0.016 to 0.033); and residence in NWU vs AUA (*β*=0.403 to 0.442 p=0.037 to 0.052) while controlling for covariates.

**Table 2.** Results of linear regression examining associations between square root-transformed sleep apnea indices and participant characteristics (n=319).

| Term | AHI 3% |  |  |  | AHI 4% |  |  |  | ODI 4% |  |  |  |
| --- | --- | --- | --- | --- | --- | --- | --- | --- | --- | --- | --- | --- |
| | $\hat{\beta}$ | 2.5th | 97.5th | p | $\hat{\beta}$ | 2.5th | 97.5th | p | $\hat{\beta}$ | 2.5th | 97.5th | p |
| Age | 0.030 | 0.012 | 0.048 | **0.002** | 0.030 | 0.012 | 0.049 | **0.001** | 0.030 | 0.011 | 0.048 | **0.002** |
| Sex, Female (Ref=Male) | -0.884 | -1.224 | -0.543 | **6.30E-07** | -0.949 | -1.295 | -0.603 | **1.52E-07** | -0.979 | -1.328 | -0.631 | **7.99E-08** |
| CREBRF genotype | 0.117 | -0.104 | 0.338 | 0.302 | 0.123 | -0.102 | 0.348 | 0.285 | 0.122 | -0.105 | 0.348 | 0.293 |
| BMI | 0.124 | 0.099 | 0.150 | **9.35E-19** | 0.136 | 0.109 | 0.162 | **6.16E-21** | 0.134 | 0.108 | 0.161 | **2.32E-20** |
| Abdominal-hip circumference ratio | 2.923 | 0.330 | 5.515 | **0.028** | 2.875 | 0.237 | 5.512 | **0.033** | 3.291 | 0.632 | 5.950 | **0.016** |
| Pre-Type 2 Diabetes (Ref=No diabetes) | 0.238 | -0.253 | 0.730 | 0.343 | 0.224 | -0.275 | 0.724 | 0.379 | 0.223 | -0.281 | 0.727 | 0.387 |
| Type 2 Diabetes (Ref=No diabetes) | 0.024 | -0.509 | 0.557 | 0.929 | 0.037 | -0.505 | 0.579 | 0.894 | 0.021 | -0.526 | 0.567 | 0.941 |
| Hypertension (Ref=No hypertension) | 0.156 | -0.190 | 0.503 | 0.377 | 0.122 | -0.231 | 0.475 | 0.498 | 0.148 | -0.207 | 0.504 | 0.415 |
| Asthma (Ref=No asthma) | 0.144 | -0.626 | 0.913 | 0.714 | -0.064 | -0.846 | 0.719 | 0.874 | -0.103 | -0.892 | 0.686 | 0.798 |
| Census region, NWU (Ref=AUA) | 0.403 | -0.001 | 0.807 | <u>0.052</u> | 0.427 | 0.016 | 0.839 | **0.042** | 0.442 | 0.028 | 0.857 | **0.037** |
| Census region, ROU (Ref=AUA) | -0.021 | -0.434 | 0.391 | 0.919 | 0.029 | -0.391 | 0.448 | 0.894 | 0.025 | -0.398 | 0.448 | 0.907 |
| Partnered (Ref=Not partnered) | -0.123 | -0.527 | 0.280 | 0.550 | -0.158 | -0.569 | 0.253 | 0.452 | -0.178 | -0.592 | 0.236 | 0.400 |
| Socioeconomic resources | 0.013 | -0.027 | 0.053 | 0.511 | 0.028 | -0.013 | 0.069 | 0.176 | 0.031 | -0.010 | 0.072 | 0.144 |
| Education | -0.020 | -0.080 | 0.040 | 0.521 | -0.016 | -0.077 | 0.045 | 0.610 | -0.016 | -0.077 | 0.046 | 0.612 |
| Cigarette use (Ref=No) | -0.179 | -0.503 | 0.146 | 0.282 | -0.086 | -0.416 | 0.245 | 0.611 | -0.091 | -0.424 | 0.242 | 0.593 |
| Alcohol use (Ref=No) | 0.174 | -0.423 | 0.772 | 0.568 | 0.193 | -0.415 | 0.800 | 0.534 | 0.222 | -0.391 | 0.834 | 0.479 |

$\hat{\beta}$ = regression estimate for square root transformed dependent variable (sleep apnea measure).
2.5<sup>th</sup> = lower bound of 95% confidence interval for $\hat{\beta}$ ; 97.5<sup>th</sup> = upper bound of 95% confidence interval for $\hat{\beta}$ .
Bolded p-values indicate p<0.05, underlined p-values indicate p<0.1.
AHI = apnea hypopnea index; ODI = oxygen desaturation index; CREBRF rs373863828 genotype modeled additively based on number of copies of minor allele; BMI = body mass index (kg/m<sup>2</sup>); Pre-T2DM and T2DM = Type 2 diabetes status determined based on any of the following: current use of diabetes medication, Hemoglobin A1c (HbA1c) (5.7-6.4%, pre-diabetes; >6.4%, diabetes), fasting blood glucose (FBG; 100-125, pre-diabetes; ≥126, diabetes), and/or oral glucose tolerance testing based on the American Diabetes Association criteria; Hypertension, defined as an average blood pressure value ≥140/90 mmHg and/or current hypertension medication use; Asthma, based on self-report history of asthma diagnosis; Partnered = currently married or cohabitating (vs. not partnered = never married, separated, divorced, or widowed); Socioeconomic resources measured based on the number of household assets as a measure of material lifestyle score/socioeconomic position; Education, self-reported years of education; Smoking, defined based on self-report of current use of cigarettes, cigars, or pipes; Alcohol, defined based on self-report of consumption of alcohol in the past 12 months.

## 4. DISCUSSION

Our study highlights a high prevalence of sleep apnea in this sample of adults from Samoa, emphasizing the urgency of addressing this public health issue. We found that 87.9% and 54.9% of the sample had sleep apnea with >5 or ≥15 events/hr, respectively, based on the AHI 3% metric. This exceeds values reported in a previous study^2^, which *estimated* (based only on population similarity to other areas with similar BMI and geographical proximity) lower expected prevalence rates in Samoa of 33.6% and 14.7%, respectively, by similar definition.^2^

Beyond this work, only two other research papers have focused on sleep health in Samoa. The first examined the relationship between sleep apnea treatment and blood pressure reduction among adults from Samoa.^13^ Specifically, a 2009 study found that sleep apnea treatment by CPAP resulted in a 12.9/10.5 mm Hg reduction in BP over a 6-month period in the overall group, with an even greater reduction of 21.5/13.1 mm Hg in the baseline hypertensive group.^13^ As mentioned above as an underlying motivation for our follow up study, the second paper was from our own group and characterized self-reported daytime sleepiness and insomnia in the same sample of adults from Samoa as this study and found a generally higher sample prevalence of excessive daytime sleepiness, but lower sample prevalence of insomnia compared with individuals from high-income countries.^15^

To our knowledge there are no other studies focused on sleep apnea in Samoa. Direct comparisons with other populations need to be done cautiously due to the influences of age, sex, BMI, T2DM distributions, social determinants of health, and cultural differences on sleep apnea measures as well as the influences of sleep apnea measurement differences themselves. However, to put these findings in the context of existing research, the frequency of AHI 3% potentially actionable (>5 events/hr) and clinically actionable (≥15 events/hr) sleep apnea (AHI 3%) of 87.9% and 54.9%, respectively in the current study was higher than in the US-based Multi-Ethnic Study of Atherosclerosis (MESA) which measured sleep apnea by similar definition, but using polysomnography, across a variety of racial/ethnic groups including White (62.9% and 30.3%, respectively), Black (63.5% and 32.4%, respectively), Hispanic (71.5% and 38.2%, respectively), and Chinese (66.4% and 39.4%, respectively) participants.^26^ Further, when compared to a US-based sample of Mexican American adults from Starr County, Texas, who are very different from Samoans in terms of environment and culture, but who shared similarities in terms of age range, technology (WatchPAT), and a high prevalence of T2DM, the Samoan sample exhibited a higher frequency of moderate or severe sleep apnea.^27^ For instance, within the subgroup of Starr County participants with T2DM, 52.5% of men and 48.1% of women had clinically actionable sleep apnea.^27^ In the Samoan sample, where only 36.5% had T2DM, the rates were either higher (in men) or similar (in women), with 61.7% of men and 48.9% of women having clinically actionable sleep apnea. Finally, in another high risk group, African-Americans living in the southeast US, (the Jackson Heart Sleep Study; mean age 63 years, 66% female, mean BMI 32 kg/m^2^), the prevalence of AHI 4% >5 or ≥15 was 55.1% and 23.6%, respectively, measured via an in-home Embletta-Gold sleep apnea test.^28^ These figures were also lower than the rates observed in this study, which documented rates of 68.5% and 31.5%, respectively.

Compared with other studies, we identified similar associations between AHI and ODI measures and participant factors such as male sex^29,30^, higher BMI^30^, higher age^30^, an abdominal-hip circumference ratio.^31^ In bivariate analyses, we also identified associations that persisted from the literature, including those with higher AHI/ODI measures and more socioeconomic resources^32^, less physical activity^33^, and the presence of hypertension.^34^ Of note, we also identified associations with *CREBRF* genotype – though this association did not persist when controlling for BMI. Interestingly, we identified no associations between sleep apnea measures and T2DM status in this sample despite strong associations reported in prior work^35,36^; additional research is warranted to better understand this observation. We also did not identify associations between sleep apnea and unique environmental/lifestyle variables including sleep surface, season of assessment, or room/sleep surface sharing. Given the paucity of research concerning Pacific Islanders, coupled with the alarming evidence (presented above) that points toward a disproportionate burden of sleep issues in this group, it is evident that prioritizing sleep health research within this historically underrepresented group is imperative. This will enable a more comprehensive understanding of how age, sex, BMI, T2DM, social determinants of health, and cultural factors collectively impact sleep apnea measurements and their implications for cardiometabolic health.

While there are many strengths to this study, including that it provides one of the first examinations of sleep apnea in Samoa using clinically reliable devices, the sample ascertainment bias that resulted from intentional oversampling of the *CREBRF* obesity-risk allele potentially reduces the generalizability of our findings as a product of an upwardly biased sample mean BMI. Specifically, the frequency of the *CREBRF* minor allele (A) in our sample was 0.561 compared with a lower frequency of 0.259 in the general Samoan population.^17^ The sample mean BMI of the present study, 35.5 kg/m^2^, was higher than that from the larger more generally representative (but earlier) 2010 sample of 33.5 kg/m^2^.^17^ Of note, the participants for this study were recruited through the earlier 2010 study, so this increase in BMI is likely a product of not only *CREBRF* obesity-risk allele overrepresentation, but also year-on-year effects associated with aging and nutritional transition. However, even after adjusting for CREBRF-related sample ascertainment bias (Code S1), the sleep apnea category distributions shift by only 1-2% (Table S3). Further, we don’t believe that the rates of sleep apnea in this sample are fully explained by obesity. In a sub-analysis of individuals with a BMI <30 kg/m^2^ (n=73), a substantial 27.3% exhibited an AHI 3% ≥15 events/hr. This underscores the need for further research to better understand the role of not just body size, but also other factors on the development of this condition, as well as on the generalizability of findings.

Given the distribution of sleep apnea data in the study sample, there is a compelling need for further research dedicated to confirming these finding and exploring sleep apnea subtypes and endotypes. Most immediately, however, these data suggest a need for public health planning and policies geared toward the diagnosis and provision of accessible, cost-effective sleep apnea treatment options in Samoa. With the closure of a local sleep clinic in 2020 and a reported lack of CPAP machines locally, individuals suspected of having sleep apnea must currently seek screening and treatment in New Zealand or Australia. Prioritizing the development of healthcare infrastructure specifically focused on sleep-related issues is imperative. A concerted effort, integrating public health initiatives, improved medical accessibility, and dedicated research, stands as a pivotal pathway to mitigate the impact of sleep apnea in Samoa and foster a healthier future.

## DECLARATIONS

## Supporting information

AdditionalFile

## Data Availability

Data are available under dbGAP accession number phs000914.v1.p1

https://www.ncbi.nlm.nih.gov/projects/gap/cgi-bin/study.cgi?study_id=phs000914.v1.p1

## Acknowledgements

We would like to thank the participants for their involvement in this research as well as the local village authorities, the Samoa Ministry of Health, the Samoa Bureau of Statistics, and the Ministry of Women, Community and Social Development for their support of this work. A special *fa’afetai tele lava* to our research assistants – Melania Selu, Vaimoana Lupematisila, Folla Unasa, and Lupesina Vesi.

## Ethical Approval

This study was approved by the Institutional Review Boards (IRB) at Yale and Brown Universities, with Yale University serving as the IRB of record (1604017547). Data analysis activities at the University of Pittsburgh were reviewed by their IRB and were determined to be exempt (PRO16040077) based on their receipt of only deidentified data. Activities of the Sleep Reading Center were approved by Mass General Brigham (formerly Partners Healthcare). The study was also approved by the Health Research Committee of the Samoa Ministry of Health.

## Consent to Participate

Written informed consent was obtained from all participants prior to enrollment.

## Availability of Data

dbGAP accession #phs000914.v1.p1.

## Authors’ Roles

LWH and NLH developed the premise for this paper. LWH performed the analyses with guidance from DEW. LWH wrote the first draft of the manuscript, and critically revised based on co-author feedback. NLH supervised all aspects of the project. AP was responsible for overseeing data collection activities with supervision from NLH and STM. SR and BC provided oversight of sleep data acquisition. All authors contributed to the interpretation of the data and results. All authors reviewed, critically revised, and approved the final manuscript. All authors agree to be accountable for all aspects of the work.

