## AdditionalFile for "Characterization of sleep apnea among a sample of adults from Samoa"

***Supplementary Material***

Lacey W. Heinsberg, PhD, RN*; Alysa Pomer, PhD;
Brian E. Cade, PhD; Jenna C. Carlson, PhD;
Take Naseri, MBBS, MPH; Muagututia Sefuiva Reupena, MS;
Satupa'itea Viali, MBChB, MPH; Daniel E. Weeks, PhD;
Stephen T. McGarvey, PhD, MPH;
Susan Redline, MD; Nicola L. Hawley, PhD

*Corresponding author
Department of Human Genetics
School of Public Health
University of Pittsburgh


**TABLE OF CONTENTS**


| Table/Figure/Section | Title | Page |
|---|---|---|
| Abstract | *Translated Abstract (Samoan)* | 2 |
| Methods S1 | *Expanded methods: WatchPAT data cleaning and quality control* | 3 |
| Code S1 | R code used to adjust for non-representational sampling and sample enrichment for the *CREBRF* obesity-risk allele. | 4 |
| Table S1 | Comparison of sample characteristics between overall sample and subset with WatchPAT data. | 6 |
| Table S2 | Sleep apnea categories by AHI 3%, AHI 4%, and ODI 4% across supine/non-supine positions and REM and non-REM sleep cycles. | 7 |
| Table S3 | Comparison of observed sample sleep apnea category frequencies and *CREBRF*-adjusted prevalence estimates. | 8 |
| Table S4 | Percent time spent <90% oxygen saturation, broken down by sleep apnea category. | 9 |
| Figure S1 | Sina with violin plots of sleep apnea measures by categorical participant characteristics. | 10 |
| Figure S2 | Sina with violin plots of sleep apnea measures by additional sleep environmental factors. | 12 |
| Figure S3 | Rain cloud plots describing sleep apnea measures more detailed age group and sex. | 13 |
| Figure S4 | Scatterplots of sleep apnea measures by continuous level participant characteristics. | 14 |
| Table S5 | Results of linear regression examining associations of square root transformed sleep apnea measures with *CRBERF* genotype while controlling for age and sex (Table S3a), *CREBRF* genotype while controlling for age, sex, and BMI (Table S3b), and *CREBRF* genotype while controlling for age, sex, and T2DM (Table S3c). | 15 |
| Figure S5 | Mosaic plot depicting the proportional distribution of sleep apnea categories by categorical BMI and sex. | 16 |
| Figure S6 | Mosaic plot depicting the proportional distribution of sleep apnea categories by categorical BMI and categorical age. | 17 |
| Figure S7 | Mosaic plot depicting the proportional distribution of sleep apnea categories by categorical age and sex. | 18 |
| Figure S8 | Mosaic plot depicting the proportional distribution of sleep apnea categories by *CREBRF* rs373863828 genotype and sex. | 19 |

**Translated Abstract**

*A practice of our research group is to make an abstract that has been translated to Samoan available. At this time, only English language text is allowed by medRxiv. A Samoan translation is available upon request from.*

### SAMOA WATCHPAT

**Methods S1.** WatchPAT data cleaning and quality control: Data collection was attempted in 391 participants. Data were loaded from the WatchPAT device to a secure project computer and transferred to Brigham and Women's Hospital in Boston, USA via secure file transfer daily. Studies were initially scored using the device's automated zzzPAT software, editing events and sleep stages using an approach described by Zhang et al.[1] The algorithm was initially used to detect events associated with 4% desaturation criteria and then manually annotated such that events in non-REM sleep that were not accompanied by both a reciprocal decrease in the peripheral arterial tonometry signal and increase in heart rate were deleted. The AHI 3% was then calculated by retaining events that were identified to have a greater than or equal to 3% desaturation and less than a 4% desaturation and accompanied by both reciprocal peripheral arterial tonometry and heart rate changes. Any event that occurred during a position change was deleted. Sleep stages were automatically analyzed and edited if they did not show stage-specific patterns of heart rate, oxygen saturation, and peripheral arterial amplitude. Studies required a minimal of 3.7 hours of data on the oximetry and peripheral artery tonometry channels to be included. Post-QC, 330 participants were included for analyses.

**Code S1.** R code used to adjust for non-representational sampling and sample enrichment for the *CREBRF* obesity-risk allele.

```
# (1) Calculate expected genotype frequencies in the general population
# assuming a MAF p = 0.259
p <- 0.259
q <- 1 - p

freq_GG <- q^2
freq_AG <- 2 * p * q
freq_AA <- p^2

freq_GG

## [1] 0.549081

freq_AG

## [1] 0.383838

freq_AA

## [1] 0.067081

# (2) Calculate observed genotype frequencies in the sample
# 2a. Calculate the total number of participants of each genotype
total_GG <- sum(df$genotype_code == "GG")
total_AG <- sum(df$genotype_code == "AG")
total_AA <- sum(df$genotype_code == "AA")

total_GG

## [1] 145

total_AG

## [1] 139

total_AA

## [1] 46

# 2b. Calculate the frequency by dividing genotype-specific counts by total
# participant count (n=330)
freq_GG_observed <- total_GG/dim(df)[1]
freq_AG_observed <- total_AG/dim(df)[1]
freq_AA_observed <- total_AA/dim(df)[1]

freq_GG_observed

## [1] 0.4393939

freq_AG_observed

## [1] 0.4212121

freq_AA_observed

## [1] 0.1393939

# (3) Calculate weights for each genotype in our sample by dividing the
# expected (population) genotype frequency with the observed (sample) genotype
# frequency 3a. Calculate weights
weight_GG <- freq_GG/freq_GG_observed
weight_AG <- freq_AG/freq_AG_observed
weight_AA <- freq_AA/freq_AA_observed

weight_GG
```

```
## [1] 1.249633

weight_AG

## [1] 0.9112701

weight_AA

## [1] 0.4812333

# 3b. Add the weight variable to data frame
df <- df %>%
    mutate(weight = case_when(genotype_code == "GG" ~ weight_GG, genotype_code ==
        "AG" ~ weight_AG, genotype_code == "AA" ~ weight_AA))

# (4) Adjust counts of the categorical AHI variable (cahi_3p) based on the step 3 weights
counts <- df %>%
    group_by(cahi_3p) %>%
    summarise(weighted_count = sum(weight), raw_count = n()) %>%
    ungroup

counts

## # A tibble: 4 × 2
##   cahi_3p weighted_count  raw_count
##   <fct>            <dbl>      <int>
## 1 None              40.3         40
## 2 Mild             112.         109
## 3 Moderate         116.         114
## 4 Severe            61.7         67
```

SAMOA WATCHPAT

**Table S1.** Comparison of sample characteristics between overall sample and subset with WatchPAT data.

| Characteristic | Overall (n = 519) | Subset (n = 330) | p-value[4] |
|---|---|---|---|
| Age (years)[1] | 52.2 (10.0) | 52.0 (9.9) | 0.776 |
| Sex[2] | | | |
| *Male* | 233 (44.9) | 154 (46.7) | 0.613 |
| *Female* | 286 (55.1) | 176 (53.3) | |
| BMI (kg/m²)[1] | 35.8 (7.7) | 35.5 (7.5) | 0.579 |
| *Missing* | 16 | 1 | |
| Abdominal-hip ratio[1] | 0.98 (0.07) | 0.98 (0.07) | 0.999 |
| *Missing* | 16 | 2 | |
| Type 2 Diabetes[2] | | | |
| *None* | 61 (12.1) | 40 (12.2) | 0.999 |
| *Pre-diabetes* | 259 (51.5) | 169 (51.5) | |
| *Diabetes* | 183 (36.4) | 119 (36.3) | |
| *Missing* | 16 | 2 | |
| Hypertension[2] | | | |
| *No* | 324 (64.9) | 215 (66.6) | 0.630 |
| *Yes* | 175 (35.1) | 108 (33.4) | |
| *Missing* | 20 | 7 | |
| Asthma[2] | | | |
| *No* | 494 (95.6) | 316 (95.8) | 0.886 |
| *Yes* | 23 (4.4) | 14 (4.2) | |
| *Missing* | 2 | 0 | |
| *CREBRF* rs373863828 genotype[2] | | | |
| *GG* | 224 (43.2) | 145 (43.9) | 0.254 |
| *AG* | 201 (38.7) | 139 (42.1) | |
| *AA* | 94 (18.1) | 46 (13.9) | |
| Census Region[2] | | | |
| *AUA* | 116 (22.4) | 68 (20.6) | 0.753 |
| *NWU* | 221 (42.6) | 139 (42.1) | |
| *ROU* | 182 (35.0) | 123 (37.3) | |
| Education (years)[1] | 11.2 (2.7) | 11.3 (2.9) | 0.610 |
| *Missing* | 1 | 0 | |
| Relationship Status[2] | | | |
| *Not partnered* | 95 (18.3) | 57 (17.3) | 0.693 |
| *Partnered* | 423 (81.7) | 273 (82.7) | |
| *Missing* | 1 | 0 | |
| Alcohol[2] | | | |
| *No* | 478 (92.1) | 305 (92.4) | 0.864 |
| *Yes* | 41 (7.9) | 25 (7.6) | |
| Smoking[2] | | | |
| *No* | 320 (61.8) | 207 (62.9) | 0.738 |
| *Yes* | 198 (38.2) | 122 (37.1) | |
| *Missing* | 1 | 1 | |
| Material Lifestyle Score (no. household assets)[1] | 8.0 (4.0) | 8.0 (4.1) | 0.999 |
| *Missing* | 3 | 2 | |
| Accelerometer-measured physical activity (minutes/day)[3] | 73.0 (82.7) | 75.3 (88.6) | 0.497 |
| *Missing* | 87 | 39 | |

[1]mean (SD); [2]n (%); [3]median (IQR); [4]Statistical testing: T-test (variables with superscript [1]), chi-square test (variables with superscript [2]), or Mann-Whitney U Test (variables with superscript [3]); AUA = Apia Urban Area; NWU = Northwest 'Upolu; ROU = Rest of 'Upolu; Hypertension, defined as an average value ≥140/90 mmHg and/or current hypertension medication use; Diabetes, based on Hemoglobin A1c values or current use of diabetes medication; Asthma, self-report history of asthma diagnosis; Smoking, defined based on self-report of current use of cigarettes, cigars, or pipes; Alcohol, defined based on self-report of consumption of alcohol in the past 12 months; Moderate-vigorous physical activity, average minutes/day measured using accelerometer.

**Table S2.** Sleep apnea categories by AHI 3%, AHI 4%, and ODI 4% across supine/non-supine positions and REM and non-REM sleep cycles, n (%).

| | All (n=316) | | | Male (n=152) | | | Female (n=164) | | |
|---|---|---|---|---|---|---|---|---|---|
| **Supine** | **AHI 3%** | **AHI 4%** | **ODI 4%** | **AHI 3%** | **AHI 4%** | **ODI 4%** | **AHI 3%** | **AHI 4%** | **ODI 4%** |
| All categories | | | | | | | | | |
| *None/minimal sleep apnea (<5 events/hr)* | 33 (10.4) | 96 (30.4) | 87 (27.5) | 10 (6.6) | 37 (24.3) | 33 (21.7) | 23 (14.0) | 59 (36.0) | 54 (32.9) |
| *Mild (5 to <15 events/hr)* | 64 (20.3) | 89 (28.2) | 91 (28.8) | 25 (16.4) | 38 (25.0) | 40 (26.3) | 39 (23.8) | 51 (31.1) | 51 (31.1) |
| *Moderate (15 to<30 events/hr)* | 92 (29.1) | 75 (23.7) | 82 (25.9) | 36 (26.7) | 36 (23.7) | 38 (25.0) | 56 (34.1) | 39 (23.8) | 44 (26.8) |
| *Severe (≥30)* | 127 (40.2) | 56 (17.7) | 56 (17.7) | 81 (53.3) | 41 (27.0) | 41 (27.0) | 46 (27.9) | 15 (9.1) | 15 (9.1) |
| Clinical categories | | | | | | | | | |
| *Potentially actionable (≥5 events/hr)* | 283 (89.6) | 220 (69.6) | 229 (72.4) | 142 (96.4) | 115 (75.7) | 119 (78.3) | 141 (85.8) | 105 (64.0) | 110 (67.0) |
| *Clinically actionable (≥15 events/hr)* | 219 (69.3) | 131 (41.4) | 138 (43.6) | 117 (80.0) | 77 (50.7) | 79 (52.0) | 102 (62.0) | 54 (32.9) | 59 (35.9) |
| | **All (n=315)** | | | **Male (n=148)** | | | **Female (n=167)** | | |
| **Non-supine** | **AHI 3%** | **AHI 4%** | **ODI 4%** | **AHI 3%** | **AHI 4%** | **ODI 4%** | **AHI 3%** | **AHI 4%** | **ODI 4%** |
| All categories | | | | | | | | | |
| *None/minimal sleep apnea (<5 events/hr)* | 66 (21.0) | 153 (48.6) | 135 (42.9) | 26 (17.6) | 74 (50.0) | 69 (46.6) | 40 (24.0) | 79 (47.3) | 66 (39.5) |
| *Mild (5 to <15 events/hr)* | 82 (26.0) | 97 (30.8) | 104 (33.0) | 37 (25.0) | 37 (25.0) | 36 (24.3) | 45 (26.9) | 60 (35.9) | 68 (40.7) |
| *Moderate (15 to<30 events/hr)* | 84 (26.7) | 38 (12.1) | 42 (13.3) | 40 (27.0) | 21 (14.2) | 23 (15.5) | 44 (26.3) | 17 (10.2) | 19 (11.4) |
| *Severe (≥30)* | 83 (26.3) | 27 (8.6) | 34 (10.8) | 45 (30.4) | 16 (10.8) | 20 (13.5) | 38 (22.8) | 11 (6.6) | 14 (8.4) |
| Clinical categories | | | | | | | | | |
| *Potentially actionable (≥5 events/hr)* | 249 (79.0) | 162 (51.5) | 180 (57.1) | 122 (82.4) | 74 (50.0) | 79 (53.3) | 127 (76.0) | 88 (52.7) | 101 (60.5) |
| *Clinically actionable (≥15 events/hr)* | 167 (53.0) | 65 (20.7) | 76 (24.1) | 85 (57.4) | 37 (25.0) | 43 (29.0) | 82 (49.1) | 28 (16.8) | 33 (19.8) |
| | **All (n=303)** | | | **Male (n=138)** | | | **Female (n=165)** | | |
| **REM** | **AHI 3%** | **AHI 4%** | **ODI 4%** | **AHI 3%** | **AHI 4%** | **ODI 4%** | **AHI 3%** | **AHI 4%** | **ODI 4%** |
| All categories | | | | | | | | | |
| *None/minimal sleep apnea (<5 events/hr)* | 20 (6.6) | 58 (19.1) | | 8 (5.8) | 28 (20.3) | | 12 (7.3) | 30 (18.2) | |
| *Mild (5 to <15 events/hr)* | 62 (20.5) | 75 (24.8) | | 29 (21.0) | 31 (22.5) | | 33 (20.0) | 44 (26.7) | |
| *Moderate (15 to<30 events/hr)* | 87 (28.7) | 76 (25.1) | | 41 (29.7) | 34 (24.6) | | 46 (27.9) | 42 (25.5) | |
| *Severe (≥30)* | 134 (44.2) | 94 (31.0) | | 60 (43.5) | 45 (32.6) | | 74 (44.8) | 49 (29.7) | |
| Clinical categories | | | | | | | | | |
| *Potentially actionable (≥5 events/hr)* | 283 (93.4) | 245 (80.9) | | 130 (94.2) | 110 (79.7) | | 153 (92.7) | 135 (81.9) | |
| *Clinically actionable (≥15 events/hr)* | 221 (72.9) | 170 (56.1) | | 101 (73.2) | 79 (57.2) | | 120 (72.7) | 91 (55.2) | |
| | **All (n=303)** | | | **Male (n=138)** | | | **Female (n=165)** | | |
| ***Non-REM*** | **AHI 3%** | **AHI 4%** | **ODI 4%** | **AHI 3%** | **AHI 4%** | **ODI 4%** | **AHI 3%** | **AHI 4%** | **ODI 4%** |
| All categories | | | | | | | | | |
| *None/minimal sleep apnea (<5 events/hr)* | 68 (22.4) | 133 (43.9) | | 25 (18.1) | 50 (36.2) | | 43 (26.1) | 83 (50.3) | |
| *Mild (5 to <15 events/hr)* | 113 (37.3) | 99 (32.7) | | 44 (31.9) | 42 (30.4) | | 69 (41.8) | 57 (34.5) | |
| *Moderate (15 to<30 events/hr)* | 78 (25.7) | 50 (16.5) | | 42 (30.4) | 33 (23.9) | | 36 (21.8) | 17 (10.3) | |
| *Severe (≥30)* | 44 (14.5) | 21 (6.9) | | 27 (19.6) | 13 (9.4) | | 17 (10.3) | 8 (4.8) | |
| Clinical categories | | | | | | | | | |
| *Potentially actionable (≥5 events/hr)* | 235 (77.5) | 170 (56.1) | | 113 (81.9) | 88 (63.7) | | 122 (73.9) | 82 (49.6) | |
| *Clinically actionable (≥15 events/hr)* | 122 (40.2) | 71 (23.4) | | 69 (50.0) | 46 (33.3) | | 53 (32.1) | 25 (15.1) | |

AHI, apnea hypopnea index; ODI, oxygen desaturation index; REM, rapid eye movement.

**Table S3.** Comparison of observed sample sleep apnea category frequencies and *CREBRF*-adjusted prevalence estimates.

| **Categorical AHI 3% (events/hr)** | Observed categorical counts, n (%) | *CREBRF*-adjusted categorical counts, n (%)[1] |
|---|---|---|
| ***All categories*** | | |
| *None/minimal sleep apnea (<5 events/hr)* | 40 (12.1) | 40.3 (12.2) |
| *Mild (5 to <15 events/hr)* | 109 (33.0) | 112 (33.9) |
| *Moderate (15 to<30 events/hr)* | 114 (34.6) | 116 (35.2) |
| *Severe (≥30)* | 67 (20.3) | 61.7 (18.7) |
| ***Clinical categories*** | | |
| *Potentially actionable (≥5 events/hr)* | 290 (87.9) | 289.7 (87.8) |
| *Clinically actionable (≥15 events/hr)* | 181 (54.9) | 177.7 (53.9) |
| **Categorical AHI 4% (events/hr)** | Observed categorical counts, n (%) | *CREBRF*-adjusted categorical counts, n (%)[1] |
| ***All categories*** | | |
| *None/minimal sleep apnea (<5 events/hr)* | 104 (31.5) | 108 (32.7) |
| *Mild (5 to <15 events/hr)* | 122 (37.0) | 123 (37.3) |
| *Moderate (15 to<30 events/hr)* | 69 (20.9) | 68.8 (20.8) |
| *Severe (≥30)* | 35 (10.6) | 30.2 (9.2) |
| ***Clinical categories*** | | |
| *Potentially actionable (≥5 events/hr)* | 226 (68.5) | 222 (67.3) |
| *Clinically actionable (≥15 events/hr)* | 104 (31.5) | 99 (30.0) |
| **Categorical ODI 4% (events/hr)** | Observed categorical counts, n (%) | *CREBRF*-adjusted categorical counts, n (%)[1] |
| **All categories** | | |
| *None/minimal sleep apnea (<5 events/hr)* | 95 (28.8) | 97.8 (29.6) |
| *Mild (5 to <15 events/hr)* | 121 (36.7) | 123.1 (37.3) |
| *Moderate (15 to<30 events/hr)* | 74 (22.4) | 73.6 (22.3) |
| *Severe (≥30)* | 40 (12.1) | 35.5 (10.8) |
| ***Clinical categories*** | | |
| *Potentially actionable (≥5 events/hr)* | 235 (71.2) | 232.2 (70.4) |
| *Clinically actionable (≥15 events/hr)* | 114 (34.5) | 109.1 (33.1) |

n=330. [1]To estimate sleep apnea category prevalence that adjusts for sample ascertainment bias (i.e., *CREBRF* obesity-risk allele oversampling) and is therefore likely more generalizable to the population, we used post-stratification calibration as shown in Code S1. This approach was similar to inverse probability weighting in that each individual observation was weighted by their chance of being in the sample. Specifically, we estimated genotype-specific weights as the observed/expected genotype frequency and then sleep apnea category counts were adjusted based on genotype. Observed sample genotype frequencies of GG=0.439, AG=0.421, and AA=0.139; expected genotype frequencies (based on population minor allele frequency of 0.259) of GG=0.549, AG=0.383, and AA=0.067; genotype specific weights of GG=1.251, AG=0.910, AA=0.482.

**Table S4.** Categorical percent time spent <90% oxygen saturation, broken down by sleep apnea category.

| **All participants (n=330)** | | **Sleep apnea category** | | | |
|---|---|---|---|---|---|
| **Hypoxia category**[1] | | None (n=40) | Mild (n=109) | Moderate (n=114) | Severe (n=67) |
| 0% | None | 31 (77.5) | 46 (42.2) | 9 (7.9) | 0 |
| >0 to <5% | Light | 9 (22.5) | 63 (57.8) | 95 (83.3) | 34 (50.7) |
| 5% to <10% | Mild | 0 | 0 | 8 (7.0) | 8 (11.9) |
| 10% to <25% | Moderate | 0 | 0 | 2 (1.8) | 19 (28.4) |
| ≥25% | Severe | 0 | 0 | 0 | 6 (9.0) |
| **Male participants (n=154)** | | **Sleep apnea category** | | | |
| **Hypoxia category**[1] | | None (n=18) | Mild (n=41) | Moderate (n=54) | Severe (n=41) |
| 0% | None | 12 (66.7) | 17 (41.5) | 7 (13.0) | 0 |
| >0 to <5% | Light | 6 (33.3) | 24 (58.5) | 44 (81.5) | 16 (39.0) |
| 5% to <10% | Mild | 0 | 0 | 3 (5.6) | 4 (9.8) |
| 10% to <25% | Moderate | 0 | 0 | 0 | 18 (43.9) |
| ≥25% | Severe | 0 | 0 | 0 | 3 (7.3) |
| **Female participants (n=176)** | | **Sleep apnea category** | | | |
| **Hypoxia category**[1] | | None (n=22) | Mild (n=68) | Moderate (n=60) | Severe (n=26) |
| 0% | None | 19 (86.4) | 29 (42.6) | 2 (3.3) | 0 |
| >0 to <5% | Light | 3 (13.6) | 39 (57.4) | 51 (85.0) | 18 (69.2) |
| 5% to <10% | Mild | 0 | 0 | 5 (8.3) | 4 (15.4) |
| 10% to <25% | Moderate | 0 | 0 | 2 (3.3) | 1 (3.8) |
| ≥25% | Severe | 0 | 0 | 0 | 3 (11.5) |

[1]Based on % time spent at <90% oxygen saturation

SAMOA WATCHPAT

**Figure S1.** Sina with violin plots of sleep apnea measures by categorical participant characteristics.

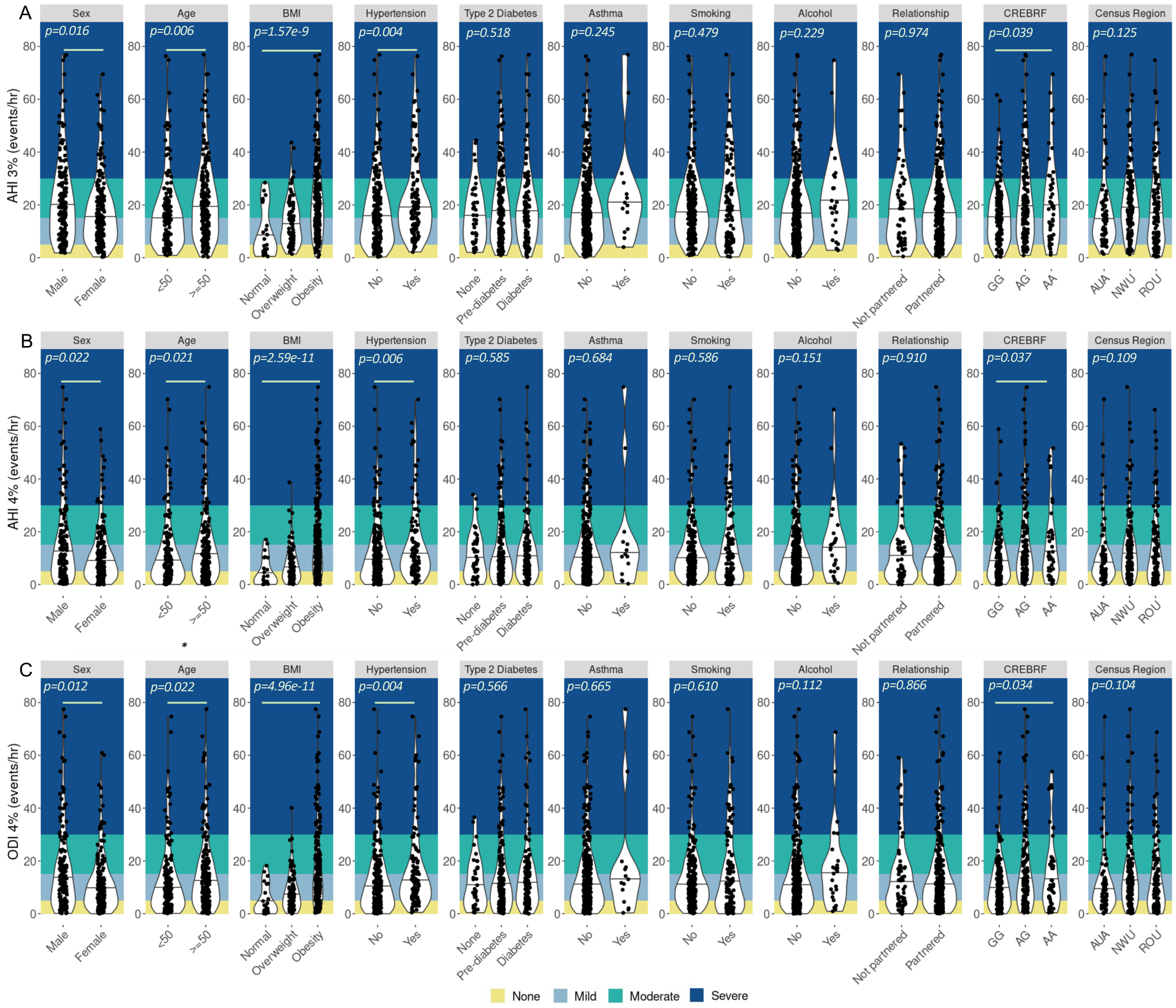


AHI = apnea hypopnea index; ODI = oxygen desaturation index; Age (years); BMI (kg/m$^2$), 'Normal' = 18-25.99 kg/m$^2$, Overweight = 26-32 kg/m$^2$, Obesity = >32 kg/m$^2$; Hypertension, defined as an average value ≥140/90 mmHg and/or current hypertension medication use; Diabetes, Type 2 diabetes status was determined based on any of the following: current use of diabetes medication,

Hemoglobin A1c (HbA1c) (5.7-6.4%, pre-diabetes; >6.4%, diabetes), fasting blood glucose (FBG; 100-125, pre-diabetes; ≥126, diabetes), and/or oral glucose tolerance testing based on the American Diabetes Association criteria; Asthma, self-report history of asthma diagnosis; Smoking, defined based on self-report of current use of cigarettes, cigars, or pipes; Alcohol, defined based on self-report of consumption of alcohol in the past 12 months; Not partnered = never married, separated, divorced, or widowed; Partnered = currently married or cohabitating; *CREBRF* genotype corresponds to rs373863828, minor allele = A; AUA = Apia Urban Area; NWU = Northwest ‘Upolu; ROU = Rest of ‘Upolu. Black horizontal line depicts subgroup median; green horizontal line indicates p-value < 0.05. Wilcox test used for binary variables, Kruskal-Wallis test used for variables with more than two levels.

**Figure S2.** Sina with violin plots of sleep apnea measures by additional sleep environmental factors.

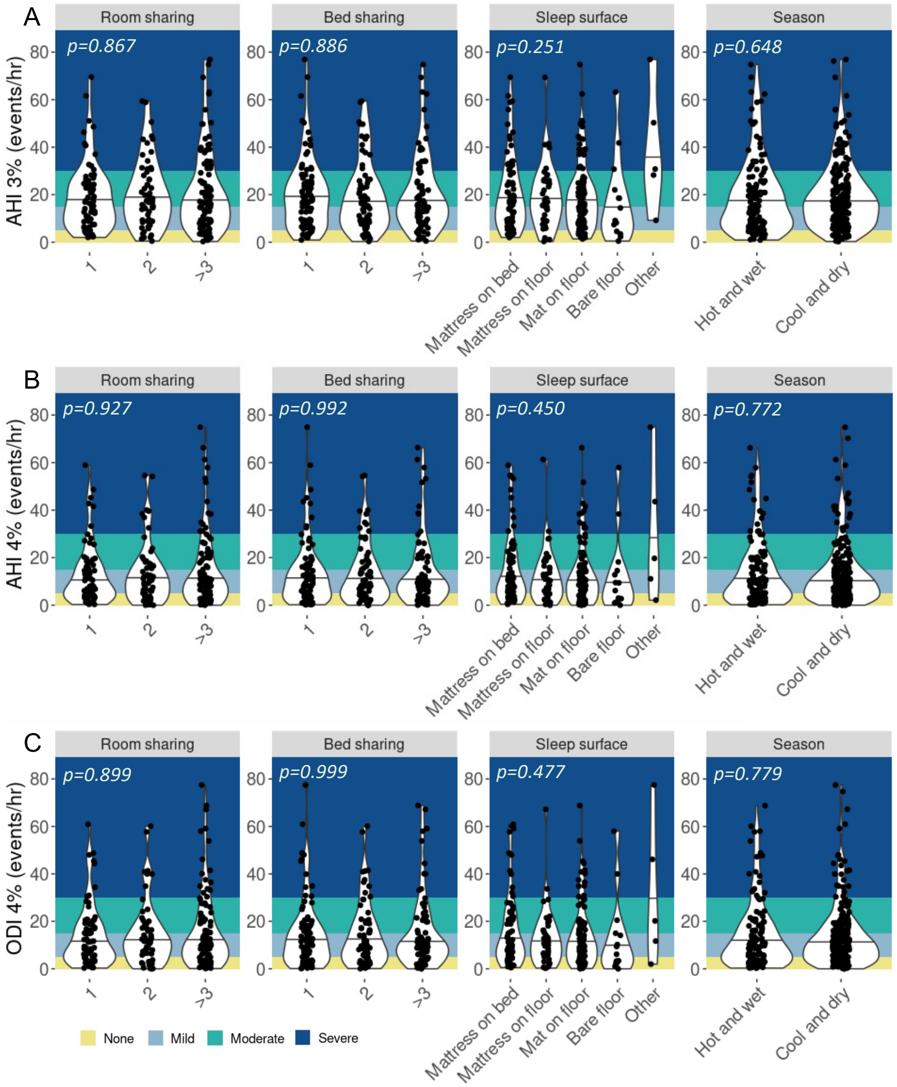


AHI = apnea hypopnea index; ODI = oxygen desaturation index; Room sharing = number of people sharing a room the night of the WatchPAT assessment, >3 category ranges from 3-9; Bed sharing = number of people sharing the sleep surface the night of the WatchPAT assessment, >3 category ranges from 3-9; Sleep surface = sleep surface the night of the WatchPAT assessment, other category includes carpet/floor/blanket/mat on sofa/sofa. Black horizontal line depicts subgroup median. Wilcox test used for binary variables, Kruskal-Wallis test used for variables with more than two levels.

**Figure S3.** Rain cloud plots describing sleep apnea measures more detailed age group and sex.

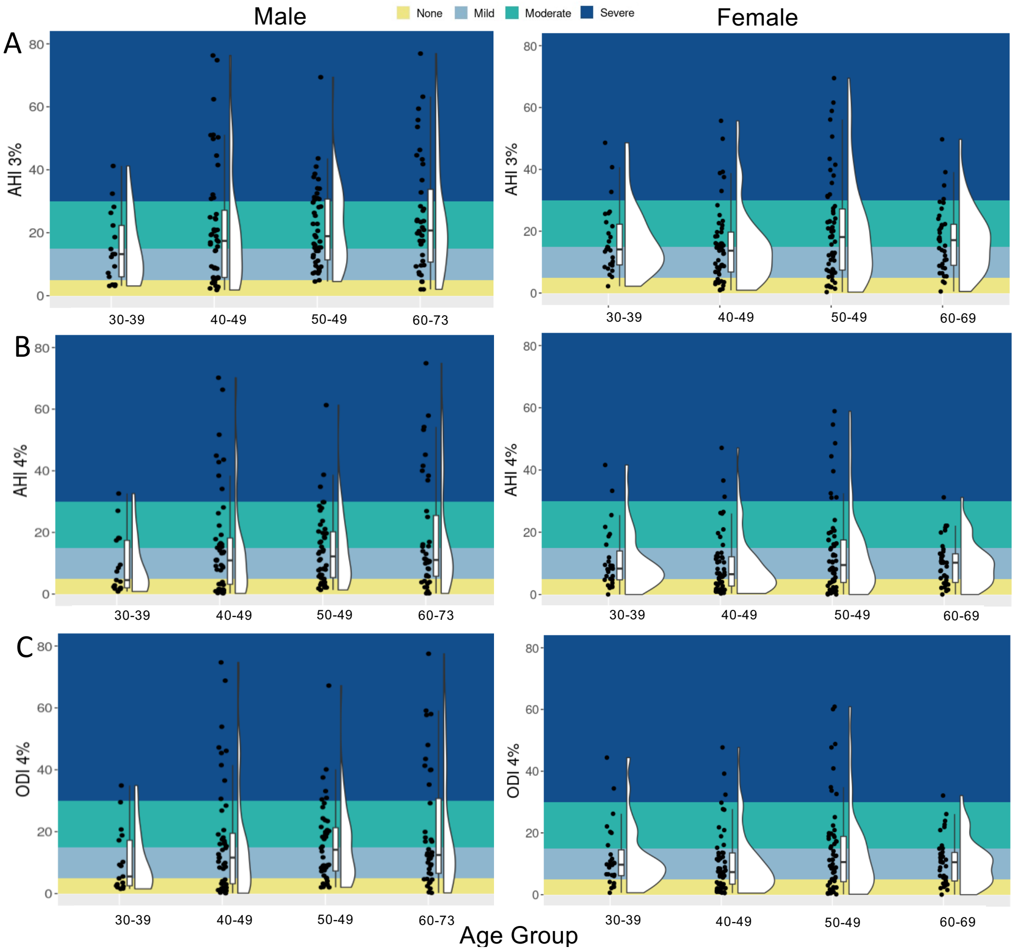


AHI = apnea hypopnea index; ODI = oxygen desaturation index. Black horizontal line depicts subgroup median.

SAMOA WATCHPAT

**Figure S4.** Scatterplots of sleep apnea measures by continuous level participant characteristics.

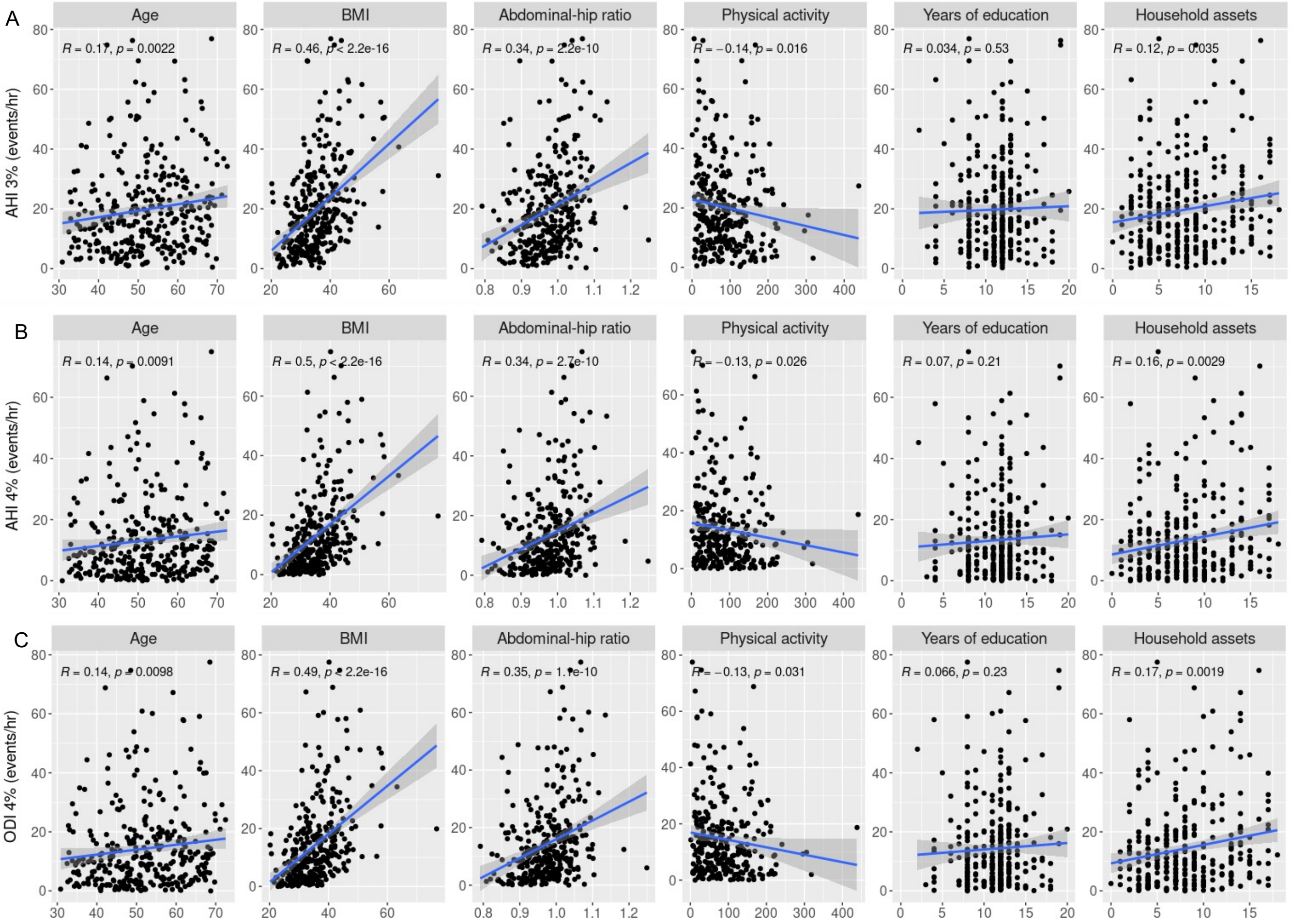


AHI = apnea hypopnea index; ODI = oxygen desaturation index; Age (years); BMI (kg/m$^2$); Physical activity, moderate-vigorous physical activity (min/day) measured using accelerometer; Household assets as a measure of material lifestyle score/socioeconomic position. R, Spearman correlation coefficient; p-value evaluated a null hypothesis of R=0; blue line indicates fitted linear regression line; gray shading indicates 95% confidence interval for regression line.

SAMOA WATCHPAT

**Table S5.** Results of linear regression examining associations of square root transformed sleep apnea measures with *CRBERF* genotype while controlling for age and sex (Table S3a), *CREBRF* genotype while controlling for age, sex, and BMI (Table S3b), and *CREBRF* genotype while controlling for age, sex, and T2DM (Table S3c).

| | AHI 3% | | | | AHI 4% | | | | ODI 4% | | | |
|---|---|---|---|---|---|---|---|---|---|---|---|---|
| **Table S3a. No covariates (n=330)** | | | | | | | | | | | | |
| **Term** | $\hat{\beta}$ | **2.5th** | **97.5th** | **p** | $\hat{\beta}$ | **2.5th** | **97.5th** | **p** | $\hat{\beta}$ | **2.5th** | **97.5th** | **p** |
| *CREBRF genotype* | 0.364 | 0.116 | 0.611 | 0.004 | 0.394 | 0.137 | 0.652 | 0.003 | 0.396 | 0.137 | 0.656 | 0.003 |
| *Age* | 0.024 | 0.007 | 0.042 | 0.007 | 0.022 | 0.004 | 0.040 | 0.020 | 0.022 | 0.003 | 0.040 | 0.022 |
| *Sex, Female (Ref=Male)* | -0.458 | -0.806 | -0.110 | 0.013 | -0.497 | -0.858 | -0.135 | 0.008 | -0.543 | -0.908 | -0.178 | 0.004 |
| **Table S3b. Controlling for BMI (n=329)** | | | | | | | | | | | | |
| | **AHI 3%** | | | | **AHI 4%** | | | | **ODI 4%** | | | |
| **Term** | $\hat{\beta}$ | **2.5th** | **97.5th** | **p** | $\hat{\beta}$ | **2.5th** | **97.5th** | **p** | $\hat{\beta}$ | **2.5th** | **97.5th** | **p** |
| *CREBRF genotype* | 0.085 | -0.127 | 0.296 | 0.433 | 0.094 | -0.122 | 0.310 | 0.394 | 0.096 | -0.123 | 0.314 | 0.391 |
| *BMI* | 0.127 | 0.106 | 0.148 | 2.03E-27 | 0.138 | 0.116 | 0.159 | 3.44E-30 | 0.138 | 0.116 | 0.159 | 1.09E-29 |
| *Age* | 0.036 | 0.021 | 0.050 | 3.49E-06 | 0.034 | 0.019 | 0.049 | 1.38E-05 | 0.034 | 0.018 | 0.049 | 1.97E-05 |
| *Sex* | -0.966 | -1.269 | -0.664 | 1.27E-09 | -1.052 | -1.361 | -0.743 | 1.12E-10 | -1.099 | -1.412 | -0.786 | 3.05E-11 |
| **Table S3c. Controlling for T2DM (n=328)** | | | | | | | | | | | | |
| | **AHI 3%** | | | | **AHI 4%** | | | | **ODI 4%** | | | |
| **Term** | $\hat{\beta}$ | **2.5th** | **97.5th** | **p** | $\hat{\beta}$ | **2.5th** | **97.5th** | **p** | $\hat{\beta}$ | **2.5th** | **97.5th** | **p** |
| *CREBRF genotype* | 0.397 | 0.144 | 0.650 | 0.002 | 0.435 | 0.172 | 0.698 | 0.028 | 0.437 | 0.172 | 0.702 | 0.001 |
| *Pre-T2DM (Ref=No diabetes)* | 0.411 | -0.148 | 0.969 | 0.151 | 0.418 | -0.163 | 0.998 | 0.159 | 0.418 | -0.168 | 1.003 | 0.163 |
| *T2DM (Ref=No diabetes)* | 0.399 | -0.201 | 0.999 | 0.193 | 0.456 | -0.167 | 1.079 | 0.152 | 0.457 | -0.171 | 1.086 | 0.155 |
| *Age* | 0.023 | 0.004 | 0.411 | 0.016 | 0.020 | 0.001 | 0.039 | 0.045 | 0.019 | 8.45E-05 | 0.386 | 0.049 |
| *Sex* | -0.475 | -0.826 | -0.124 | 0.008 | -0.522 | -0.886 | -0.157 | 0.005 | -0.567 | -0.935 | -0.200 | 0.003 |

$\hat{\beta}$ = regression estimate for square root transformed dependent variable (sleep apnea measure).

$2.5^{th}$ = lower bound of 95% confidence interval for $\hat{\beta}$; $97.5^{th}$ = upper bound of 95% confidence interval for $\hat{\beta}$.

Underlined p-values indicate p<0.05.

*CREBRF* rs373863828 genotype modeled additively based on number of copies of minor allele; BMI = body mass index (kg/m2); Pre-T2DM = pre-type 2 diabetes; T2DM = type 2 diabetes.

SAMOA WATCHPAT

**Figure S5.** Mosaic plot depicting the proportional distribution of sleep apnea categories by categorical BMI and sex.

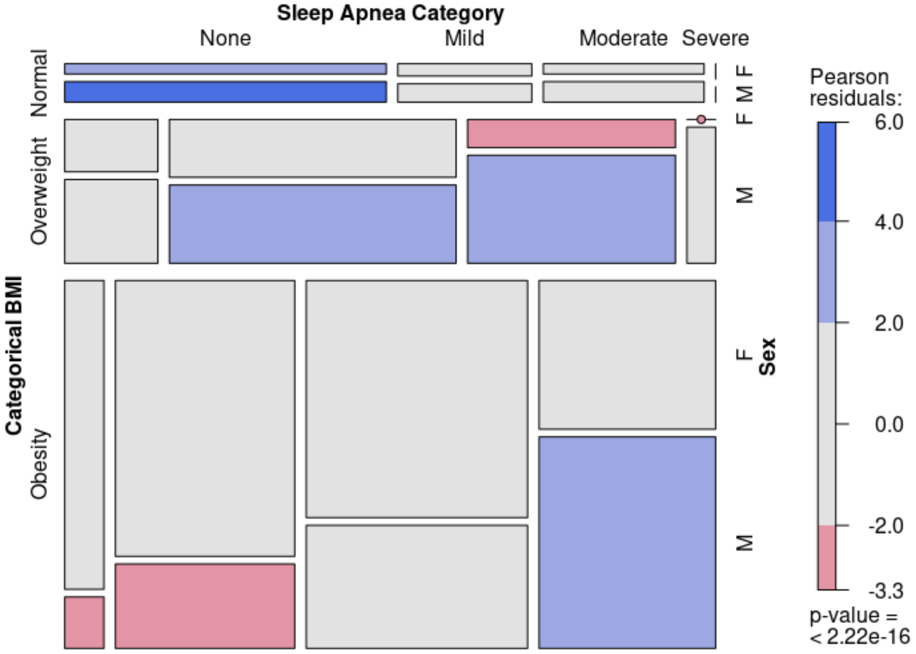


Category = AHI 3% categorized based on the number of events/hr: None (<5), Mild (5-14.9), Moderate (15-29.9), Severe (≥30). BMI, body mass index categorized: Normal (18.5 to <26 kg/m$^2$), Overweight (26 to 32 kg/m$^2$), Obesity (≥ 32 kg/m$^2$). Sex: M = male, F = female. Plot interpretation: The size of the tile is proportional to the percentage of cases in that combination of levels; assuming the three variables of interest are independent, blue shading indicates more cases than expected while red shading indicates less cases than expected (e.g., within the severe category, we see a greater number of males than expected while in the none and mild categories, we see a lower number of males than expected, suggesting a compounding of effect of sex on categorical BMI).

SAMOA WATCHPAT

**Figure S6.** Mosaic plot depicting the proportional distribution of sleep apnea categories by categorical BMI and categorical age.

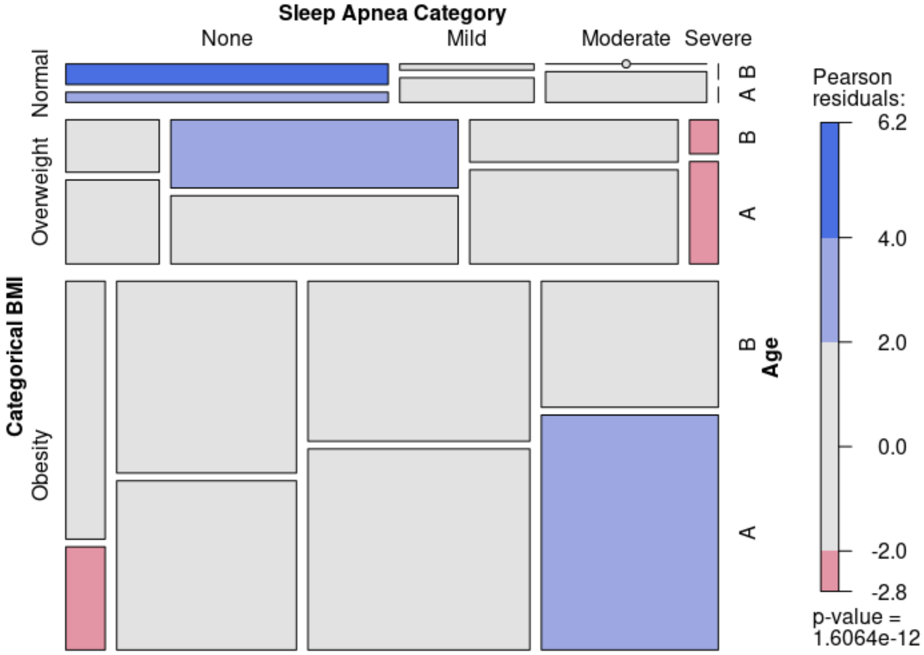


Category = AHI 3% categorized based on the number of events/hr: None (<5), Mild (5-14.9), Moderate (15-29.9), Severe (≥30). BMI, body mass index categorized: Normal (18.5 to <26 kg/m$^2$), Overweight (26 to 32 kg/m$^2$), Obesity (≥ 32 kg/m$^2$). Age: A ≥ 50 years, B < 50 years. Plot interpretation: The size of the tile is proportional to the percentage of cases in that combination of levels; assuming the three variables of interest are independent, blue shading indicates more cases than expected while red shading indicates less cases than expected.

SAMOA WATCHPAT

**Figure S7.** Mosaic plot depicting the proportional distribution of sleep apnea categories by categorical age and sex.

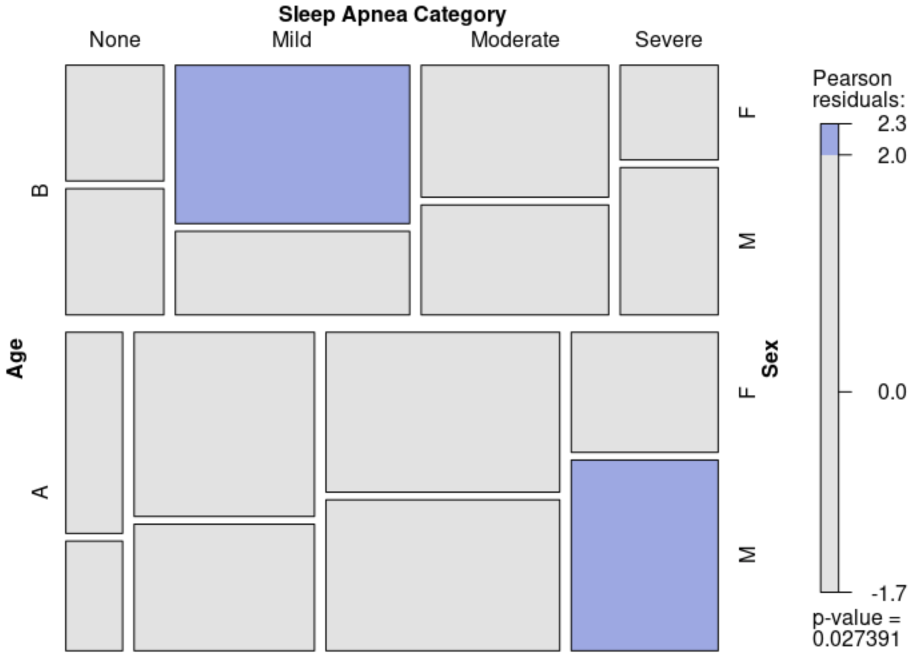


Category = AHI 3% categorized based on the number of events/hr: None (<5), Mild (5-14.9), Moderate (15-29.9), Severe (≥30). Age: A ≥ 50 years, B < 50 years. Sex: M = male, F = female. Plot interpretation: The size of the tile is proportional to the percentage of cases in that combination of levels; assuming the three variables of interest are independent, blue shading indicates more cases than expected while red shading indicates less cases than expected.

SAMOA WATCHPAT

**Figure S8.** Mosaic plot depicting the proportional distribution of sleep apnea categories by *CREBRF* rs373863828 genotype and sex.

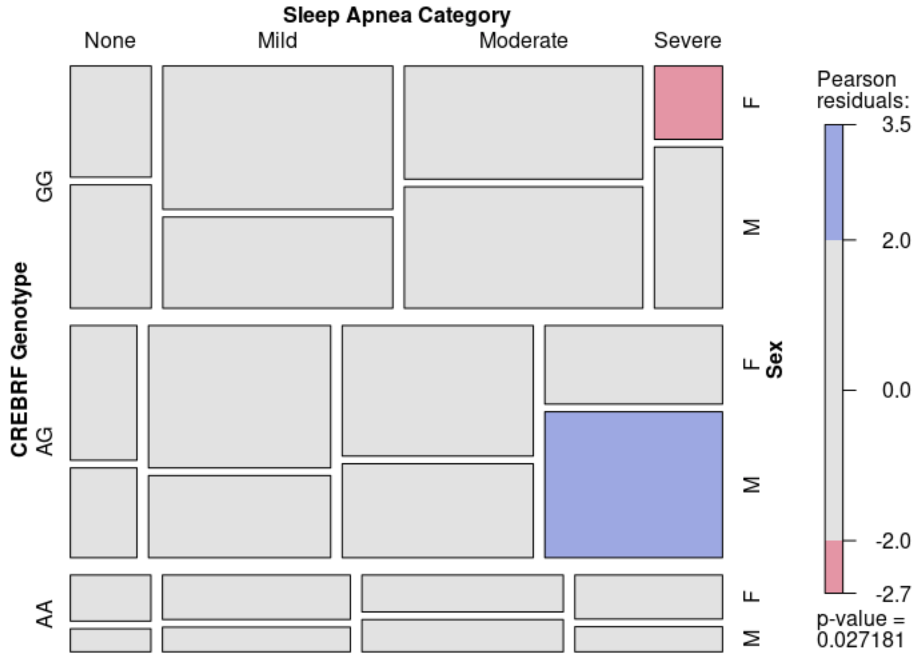


Category = AHI 3% categorized based on the number of events/hr: None (<5), Mild (5-14.9), Moderate (15-29.9), Severe (≥30). Sex: M = male, F = female. Plot interpretation: The size of the tile is proportional to the percentage of cases in that combination of levels; assuming the three variables of interest are independent, blue shading indicates more cases than expected while red shading indicates less cases than expected.
